# Dietary factors and hypertension risk in West Africa: A systematic review and meta-analysis of observational studies

**DOI:** 10.1101/2023.03.21.23287434

**Authors:** Nimisoere P. Batubo, J. Bernadette Moore, Michael A. Zulyniak

**Affiliations:** Nutritional Epidemiology Group, School of Food Science and Nutrition, University of Leeds, Leeds, LS2 9JT

**Keywords:** Hypertension, meta-analysis, West Africa, dietary factors, systematic review

## Abstract

**Background:** Contrary to North America and Europe, the prevalence of hypertension is rising in West Africa. Although diet is implicated as a contributor to this trend, nutritional guidelines in West Africa are not tailored to address this concern. This study aimed to address this limitation by investigating dietary factors common to West Africa and evaluating their association with hypertension.

**Methods:** PubMed, Scopus, Web of Science, and Medline were searched to identify studies that investigated diet and hypertension in West African adults. All meta-analyses used a generic inverse-variance random effects model, with subgroup analyses by age, BMI, and study location, and were performed in R.

**Results:** 3298 studies were identified, of which 31 (n=48,809 participants) satisfied inclusion criteria □ all cross-sectional. Meta-analyses of the association between dietary factors and hypertension included: dietary fat (OR=1.76; 95% CI:1.44,2.14; p<0.0001), red meat (OR=1.51; 95% CI: 1.04, 2.18; p = 0.03), junk-food (OR=1.41; 95% CI: 1.19, 1.67; p < 0.0001), dietary salt (OR= 1.25; 95% CI: 1.12, 1.40; p<0.0001), alcohol (OR= 1.17; 95% CI: 1.03, 1.32; p= 0.013), and ‘fruits and vegetables’ (OR=0.80; 95% CI: 0.24, 1.17; p < 0.0001). Subgroup analyses suggested that ‘fruit and vegetable’ consumption is less protective in the elderly.

**Conclusion:** High consumption of dietary salt, red meat, dietary fat, junk food, and alcohol are associated with increased odds of hypertension, whereas high fruit and vegetable appear protective. This region-specific evidence will support the development of nutritional assessment tools for clinicians, patients, and researchers aiming to reduce hypertension in West Africa.

## Introduction

Hypertension, defined as a sustained elevation of blood pressure ≥140/ 90 mmHg, remains the leading preventable risk factor for cardiovascular disease and the number one cause of death globally [1]. Approximately 40% of people aged 30-79 years have hypertension, corresponding to one in three adults (∼1.3 billion in 2019), with two-thirds of cases living in low- and middle-income countries, including African countries [2, 3]. The most recent WHO data show that the African region has the highest prevalence of hypertension (35.5%), while the Americas have the lowest (18%) [4]. Countries such as the UK, the US, and China have seen a decrease in the prevalence of hypertension, with rates declining by 6.25%, 11.38%, and 16.3%, respectively, from 2010 to 2019 [5–7], while West African countries like Nigeria have shown a consistent increase in the prevalence of hypertension. Most recent data suggest a 15.3% increase in hypertension rates in Nigeria from 2010 to 2019 [8, 9], which has coincided with an increase in heart disease, stroke, and chronic kidney disease [10, 11].

This increased trend in hypertension in Nigeria and other West African countries has been partly attributed to increased unhealthy dietary practices (consumption of a diet that is high salt/sodium, low potassium consumption, unhealthy dietary fat and oils, refined sugar, alcohol, and low ‘fruit and vegetable’) and lack of physical activity [1, 12–15]. Indeed, diet has been attributed to nearly one-third of all hypertension cases [16]. This positions diet as a major risk factor of interest in West Africa for reducing hypertension prevalence. In the UK, from 2003 to 2011, government and industry initiatives led to mean reductions in salt (-1.4 g/day) and increased fruit and vegetable (+0.2 portion/day) intake, which coincided with reductions in mean blood pressure (systolic = −2.7±0.34 mm Hg; diastolic = - 1.1±0.23 mm Hg), stroke (-42%), and ischaemic heart disease mortality (-40%) [17]. In response, Nigeria and other West African countries outlined national nutritional guidelines to combat all non-communicable diseases (NCDs), including hypertension[18, 19]; however, the translation of these guidelines into actionable advice for clinicians for combating hypertension specifically has been a challenge, possibly because (i) clinicians in Nigeria and other West African countries have not been provided with an effective strategy to provide regionally-specific dietary information to their patients, and (ii) the evidence used to inform West African diet recommendations is based on evidence weighted towards studies from non-African countries which may not be translatable or applicable to manage the contribution of diet towards the rising trend of hypertension risk in Nigeria and other West African countries.

To address this, an understanding of the dietary habits and risk of hypertension in Nigeria and other West African countries is needed to confirm current recommendations and improve current dietary assessment strategies to combat hypertension. Therefore, this study aimed to undertake a systematic review and meta-analysis to provide regionally specific evidence relating to the association between dietary factors and hypertension risk in West Africa that can be used to inform the development of nutritional assessment tools for clinicians, patients, and researchers and contribute towards reducing the prevalence of hypertension in West Africa.

## Methods

First, we performed a pilot study and evaluated the summaries of the articles we identified through our search on PubMed and that fulfilled the eligibility criteria. This step enabled us to obtain an outline of the key aspects that ought to be encompassed in the review and helped us to prepare the protocol for the systematic review and meta-analysis.

### Study design and registration

The strategy for the systematic review and meta-analysis was predefined with PROSPERO (CRD42022339736), which followed the guidelines for the Meta-analyses of observational epidemiological studies [20], and Preferred Reporting Items for Systematic review and Meta-Analysis (PRISMA) 2015 [21].

### Literature search strategy

The systematic literature search was performed using a structured PICO (population, intervention, comparison, and outcome) framework and Medical Subject Headings (MESH) indexing with key terms (and synonyms thereof). The PICO elements included the targeted West African nations (P), high dietary exposure (I), low dietary exposure (C), and hypertension/high blood pressure (O). A comprehensive search was conducted across PubMed, Ovid Medline, Scopus, and Web of Science, to identify studies published between January 2000 and February 2023. The full search strategy used for PubMed is given in **Table S1** in the supplementary material. To ensure that no relevant studies were missed, the citation lists of included studies were searched to identify additional relevant studies that meet the inclusion criteria.

### Outcome measures

Four of the most prevalent adverse outcomes of blood pressure were chosen as outcomes of interest for this review and meta-analysis. These included high systolic blood pressure, high diastolic blood pressure, high mean arterial blood pressure, and high pulse pressure.

### Study selection

Observational studies with summary data (odds ratios (OR), relative risks (RR), or hazard ratios (HRs) with corresponding 95% confidence intervals (CI), p-value, means with standard deviation or standard error) that addressed any dietary intervention on hypertension or high blood pressure in healthy West African populations with participants aged *≥*18 years were included in the study. Studies were excluded if the data were: hypertensive subjects alone, animal and cells studies, non-West African populations, non-investigative studies (such as editorials, reviews, and conference abstracts), pregnancy studies, cardiovascular diseases (such as stroke, myocardial infarction) or studies in cancer populations, studies that are not related to diet, food or nutrition, studies involving only alcohol, smoking, or biomarkers of diet. The screening of ‘title and abstract’ and ‘full text’ was performed in duplicate in DistillerSR version 2.35 [22].

### Data collection and extraction

Using a standardized form, data from each study were independently extracted by two reviewers for all variables. Dietary factors identified from the observational studies were analyzed separately. Information extracted from each study included: study details (first author, year of publication, geographic location, number of participants, number of cases, study design), demographic information related to confounding (e.g., age, sex, country, health status), anthropometric data (e.g., body mass index (BMI), methods of quantifying and measuring diet, types of measurements and the studies outcomes (blood pressure), univariable or multivariable effect estimate (RRs, HRs, or ORs including the corresponding CIs), and adjusted covariates. Where multiple analytical models were provided, effect estimates from fully adjusted were extracted. When the risk estimates for participants were reported separately for men and women in a study or for similar food groups in this same population, the ORs were combined using a fixed-effect model [23].

### Quality assessment

To assess the quality of the studies, a modified Newcastle-Ottawa Scale (NOS) adapted for cross-sectional studies was used to evaluate the study quality of cross-sectional studies of the included studies [24]. The scale assesses the selection of participants, comparability of groups, and assessment of outcomes. A total of 0-9 was assigned to each study based on the number of stars, with a higher score indicating higher quality. The risk of bias assessment was carried out independently by two researchers. Disagreements between researchers were resolved by a third researcher.

### Data synthesis and analysis

To derive summary odds ratios (ORs) and 95% confidence intervals (Cis), we applied a meta-analysis to investigate the associations of the dietary factors of the highest consumers compared with the lowest consumers using the generic inverse-variance weighted random-effects model. Using an inverse variance method, we calculated the standard errors for the logarithm odds ratio of each study. This was, in turn, considered the estimated variance of the logarithm odds ratio. The degree of variability between studies (heterogeneity) (*τ*^2^) was estimated using the DerSimonian-Laird estimator (DL) [25]. In the meta-analyses with < 5 studies, the Hartung-Knapp-Sidik-Jonkman (HKSJ) random effects model was also used. Additionally, where ≤ 3 studies were available for meta-analysis, a fixed effects model was also performed to retain power, and Bonferroni correction was applied where necessary [26–28].

The heterogeneity between studies was evaluated using Cochran’s Q statistics [29] and the *I*^2^ statistic, with uncertainty intervals, where *I*^2^>50% indicated significant statistical heterogeneity [23, 30, 31]. Subgroup analyses and meta-regression were conducted to explore potential sources of heterogeneity by pre-specified characteristics and hypertension confounders: (i) Body mass index (BMI) (a cut-off of BMI *≥* 25 kg/m^2^ was used to classify overweight/obese), (ii) mean population age, and (iii) study location. Sensitivity analysis was performed when *I*^2^>50% to estimate the influence of a single study on the overall pooled results by systematically omitting a single study and observing its influence on the overall effect size. Publication bias was done by funnel plots inspection, where ≥ 10 studies were available for a single meta-analysis. Where publication bias was visibly apparent further analyses (Egger’s regression and rank correlation test) were undertaken to estimate the effect of reporting bias on study results using the standard error of the observed outcomes as predictors, which are used to check for funnel plot asymmetry [32, 33]. The trim-and-fill method was also used to correct publication bias if detected. Where no significant effects were found, post-hoc power analyses were undertaken using fixed-effects (τ2 = 0) or random-effects (τ2>0) methods. Power *≥* 80% was considered adequate [23]. All statistical analyses were performed using meta-package (version 6.0-0) [34] in R [35].

## Results

### Literature search

The literature search and study selection processes are summarised in the PRISMA flow diagram in **Figure 1**. A total of 5883 records were identified from the initial database search (Medline =1371, PubMed=1644, Scopus =1599, Web of Science =1269). After removing duplicates (n=2585 records) and those that did not fit the inclusion criteria (n=3187), 123 articles were retrieved for full-text screening, from which 31 studies were included in the meta-analysis. Seventeen studies reported on fruit and vegetable, 10 on fruit/vegetables, 6 on junk food (fried and fast food), 4 on red meat, 5 on dietary fats (fatty food), and 8 studies reported on alcohol.

**Figure 1:**
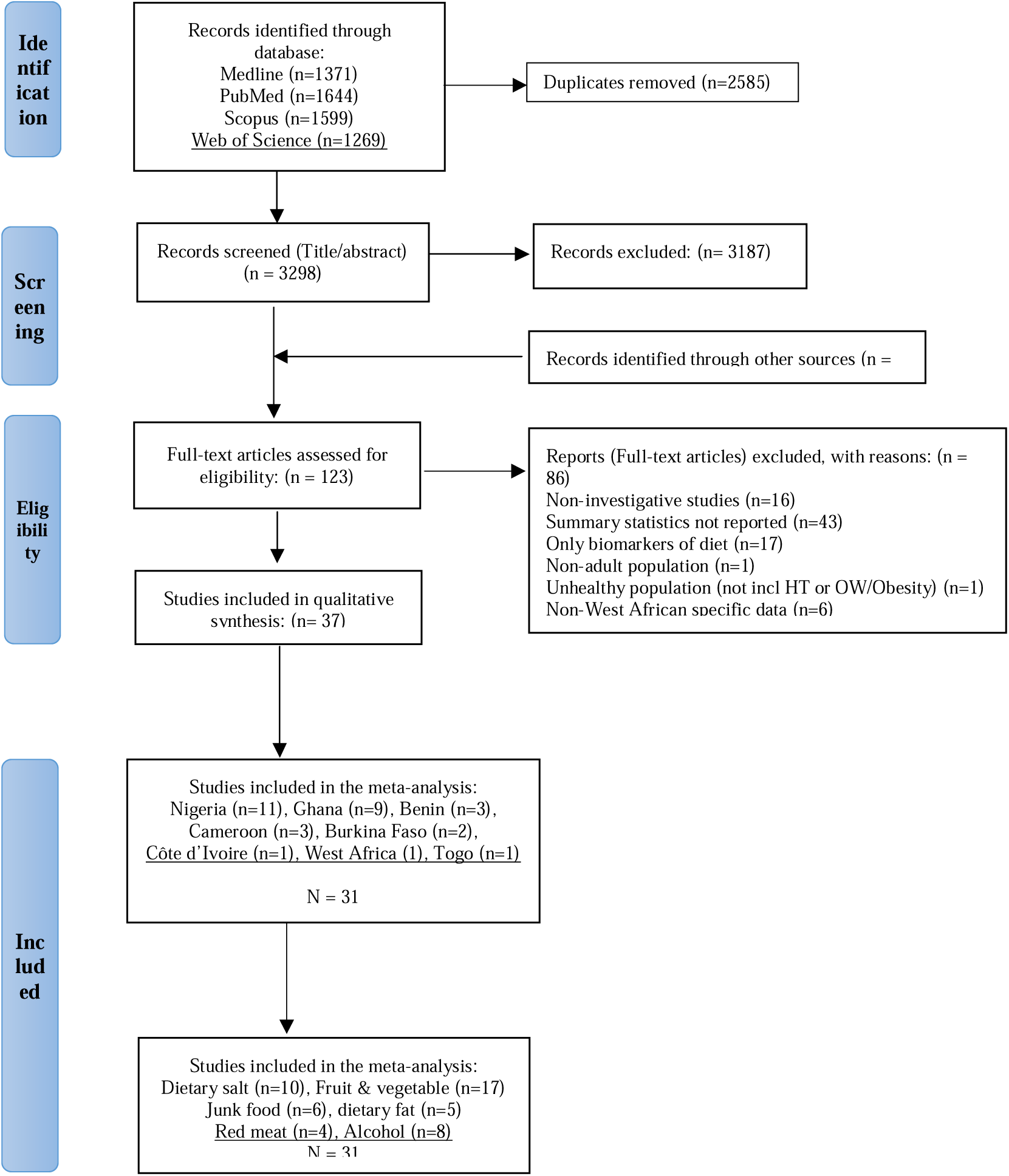
PRISMA flow diagram. Outline of identification of studies in OVID Medline, Web of Science, Scopus, and PubMed and record of the screening process and selection of articles for the systematic review and meta-analysis of the association between dietary factors and hypertension in West Africa. HT-hypertension, incl: including OW: overweight.

### Study characteristics

In total, 31 cross-sectional studies in West Africa □ including Nigeria (n=11), Ghana (n=9), Côte d’Ivoire (n=1), Benin republic (n=3), Cameroon (n=3), Burkina Faso (n=2), Togo (n=1), and 1 study from West Africa (Nigeria=1 report, Ghana= 1 report, and Burkina Faso= 1 report) □ were included in the meta-analysis (**Table S2**). Sample sizes ranged from 134 to 9,367, with a total of 48,809 participants, of which 12,898 (26.4%) were cases of hypertension. Dietary factors were assessed by the survey instrument (WHO STEPwise approach to NCD risk factor surveillance), food frequency (FFQ), or validated questionnaire.

### Data quality

The quality was assessed using a modified Newcastle-Ottawa Scale (NOS) for cross-sectional studies [24]. The criteria for allocating stars (out of a total of 9 stars) awarded to each study according to this NOS criteria can be found in **Table S3**. The 31 studies included in the meta-analysis were rated with high quality (score 7-9) and a low bias risk.

### Dietary factors associated with hypertension in West Africa

**Figure 2** illustrates the random effects model meta-analysis of the pooled estimates (odds ratio and 95% confidence interval) and heterogeneity of the dietary factors associated with hypertension when comparing high versus low consumers in West Africa. The dietary factors include fruit and vegetable, dietary salt, junk food, red meat, dietary fat, and alcohol.

**Figure 2:**
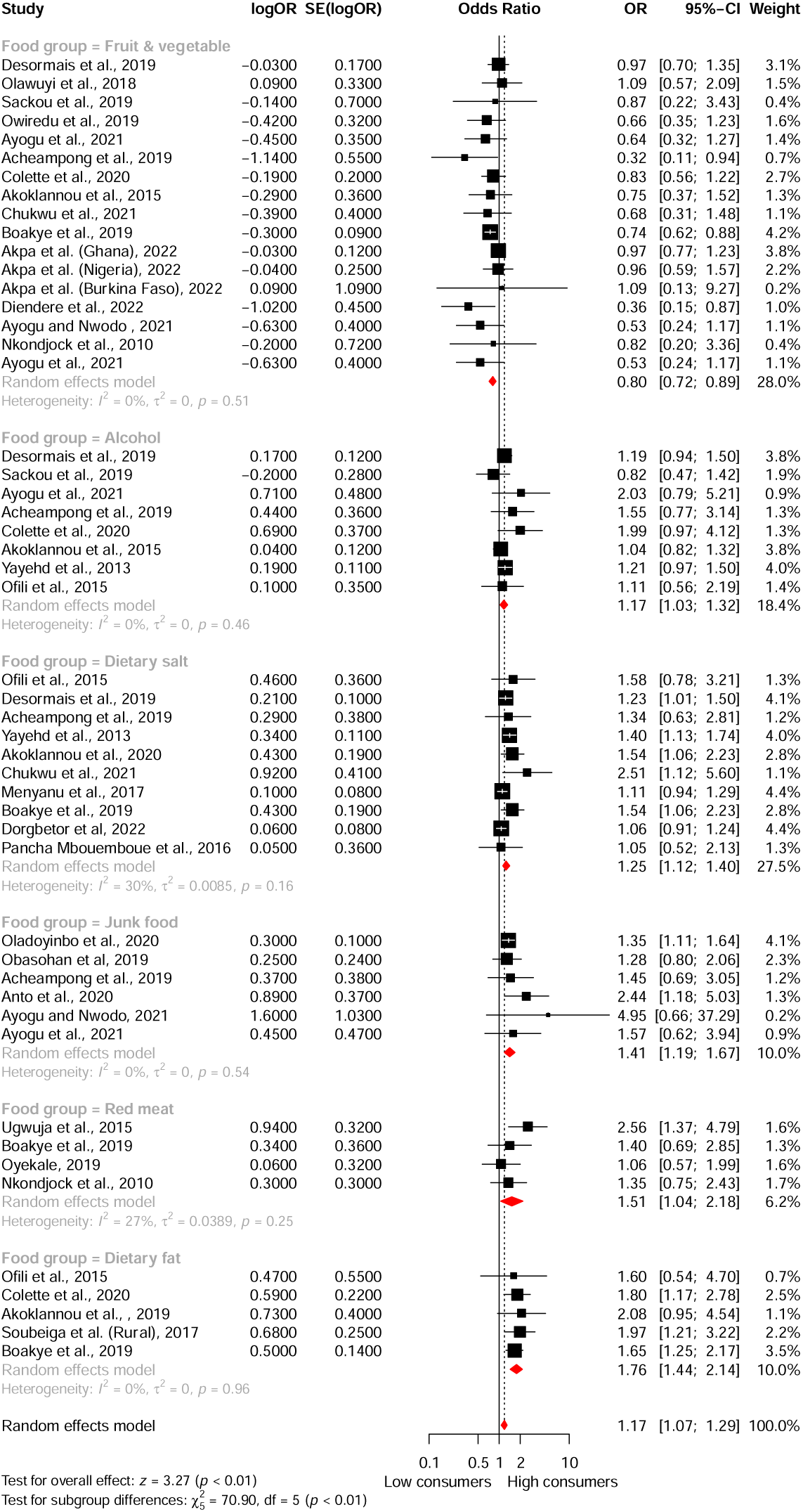
Meta-analysis of the association between dietary factors and hypertension in West Africa: Illustrating the identified dietary factors, the effect sizes, and the overall estimate of the random effects model and heterogeneity. CI: Confidence interval, logOR: Treatment effect, OR: Odds ratio, SE(logOR): Standard error.

### Fruit and vegetable consumption

Seventeen cross-sectional studies from West Africa with 19,675 participants reported on the association between fruit and vegetable consumption with the risk of hypertension □ 6,376 (32.4%) adults had hypertension. The types of fruit and vegetable investigated were not specified. Mean effect sizes ranged from 0.32 to 1.09. The overall meta-analysis suggested that consuming high amounts of fruit and vegetable was associated with 20% reduced odds of hypertension (OR=0.80; 95% CI: 0.24, 1.17; p < 0.0001, *I^2^* = 0%) compared to low consumers (**Figure 3**). No heterogeneity or potential publication bias was evident in the funnel plot (**Figure S4**), rank correlation test (p=0.17), or Egger’s regression test (p = 0.18). Subgroup analysis and meta-regression indicated that while a protective effect of fruit and vegetable consumption against hypertension was observed in all West African nations, age significantly moderated the association between fruit and vegetable consumption and hypertension. It may be less pronounced in individuals aged ≥ 50 years. However, this may be due to fewer studies in the subgroup of individuals aged ≥ 50 years. Conversely, BMI and study location did not appear to modify the association (**Table 1** and **Figures S2 and S3**). According to the results, high consumption of fruits and vegetables is associated with a 20% reduction in the odds of hypertension in West Africa. Furthermore, this correlation appears to be less significant among elderly individuals.

**Figure 3:**
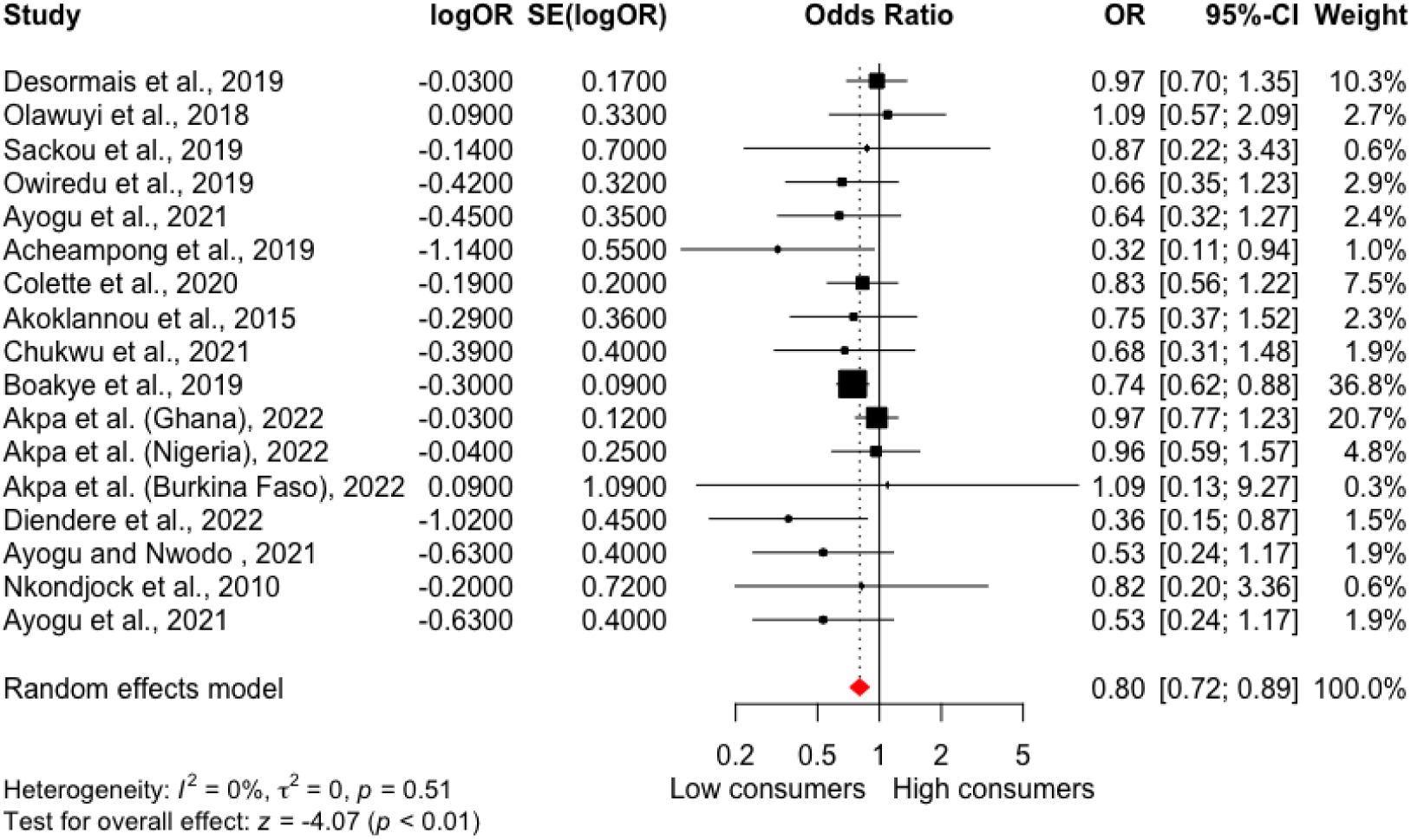
Meta-analysis of the association between fruit and vegetable consumption and hypertension in West Africa. High fruit and vegetable consumption reduces the odds of hypertension by 20% with no heterogeneity compared to low consumption. CI: Confidence interval, logOR: Treatment effect, OR: Odds ratio, SE(logOR): Standard error.

**Table 1:** Meta-regression results for moderating effects of mean age, study location, and BMI on the association between fruit and vegetable consumption and hypertension in West Africa.

| Moderator | n | Coefficient (95% CI) | * *p* -Value | * *I* <sup>2</sup> (%) | *R*<sup>2</sup> index (%) | *H*<sup>2</sup> |
| --- | --- | --- | --- | --- | --- | --- |
| **Bivariate meta-regression** |  |  |  |  |  |  |
| Mean age (years) | 17 | 0.015 (0.001, 0.030) | 0.04* | 0 | 0 | 1.00 |
| Body mass index (kg/m<sup>2</sup>) | 17 | -0.037 (-0.134, 0.059) | 0.45 | 0 | 0 | 1.00 |
| Location (Nigeria as ref.) | 17 | 0.051 (-0.245, 0.347) | 0.74 | 0 | 0 | 1.00 |
| **Multivariate meta-regression** |  |  |  |  |  |  |
| **Test of moderators** | 17 | QM (df = 3) = 1.0 | 0.80 | 0 | 0 | 1.00 |
| Mean age (years) | 17 | 0.015 (-0.016, 0.046) | 0.33 |  |  |  |
| Body mass index (kg/m<sup>2</sup>) | 17 | -0.033 (-0.195, 0.129) | 0.67 |  |  |  |
| location (Nigeria as ref.) | 17 | -0.081 (-0.690, 0.528) | 0.79 |  |  |  |

Abbreviations:
CI: Confidence interval,
*H*<sup>2</sup>: Unaccounted variability/ sampling variability
*I*<sup>2</sup>: Residual heterogeneity / unaccounted variability,
n- Number of studies,
OR-Odds ratio,
*R*<sup>2</sup>: Amount of heterogeneity accounted for.
\**I*<sup>2</sup>, and \**p*-Value related to the test of moderators.
\* Significant at 0.05

### Dietary salt consumption

A total of 10 cross-sectional studies (n=16,535 participants) reported on the association between dietary salt and hypertension risk □ 4497 (27.2%) adults had hypertension. Mean effect sizes ranged from 1.05 to 2.51. The analysis revealed that high consumption of dietary salt increased the odds of hypertension by 25% in West Africa (OR= 1.25; 95% CI: 1.12, 1.40; p<0.0001, *I^2^*=30%) compared to low consumers with moderate amount heterogeneity (**Figure 4**). No potential publication bias was evident in the funnel plot (**Figure S4**), rank correlation test (p=0.05), or Egger’s regression test (p = 0.31). Subgroup analysis and meta-regression suggested that mean age, BMI, and study location did not seem to modify the association, as shown in **Table 4 and Figures S5 and S6**. This analysis demonstrated that high dietary salt consumption increased the odds of hypertension by 25% in the West African adult population.

**Figure 4:**
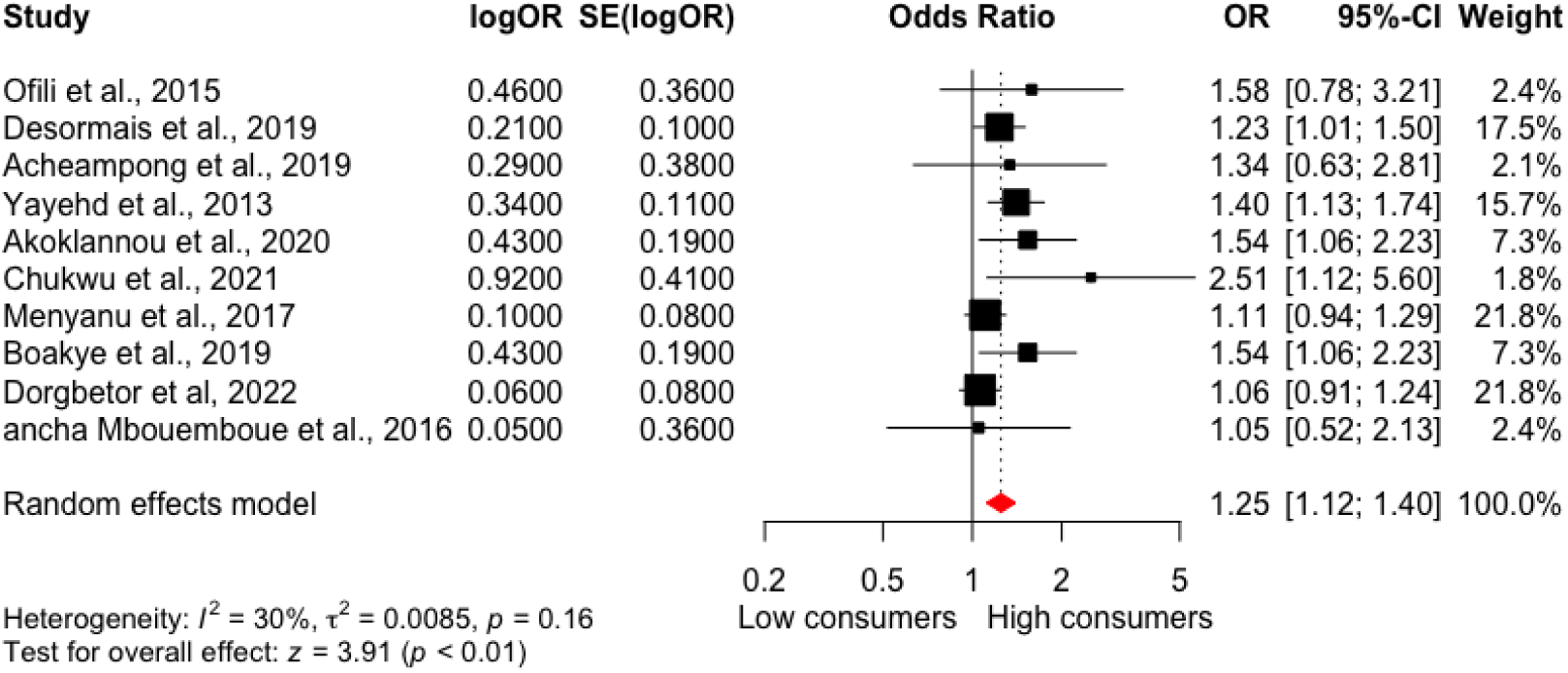
Meta-analysis of the association between dietary salt consumption and hypertension in West Africa. High dietary consumption increases the odds of hypertension by 25% with moderate heterogeneity compared to low consumption. CI-Confidence interval, logOR- Treatment effect, OR- Odds ratio, SE(logOR)- Standard error.

**Table 2:** Meta-regression results for moderating effects of mean age, study location, and BMI on the association between dietary salt consumption and hypertension in West Africa.

| Moderator | n | Coefficient (95% CI) | *p Value | *I<sup>2</sup> (%) | R<sup>2</sup> index (%) | H<sup>2</sup> |
| --- | --- | --- | --- | --- | --- | --- |
| **Bivariate meta-regression** |  |  |  |  |  |  |
| Mean age (years) | 10 | -0.004 (-0.019, 0.012) | 0.63 | 36 | 0 | 1.58 |
| Body mass index (kg/m<sup>2</sup>) | 10 | 0.022 (-0.027, 0.072) | 0.38 | 28 | 4 | 1.38 |
| Location (Nigeria as ref.) | 10 | -0.468 (-1.015, 0.079) | 0.09 | 19 | 52 | 1.23 |
| **Multivariate meta-regression** |  |  |  |  |  |  |
| **Test of moderators** | 10 | QM (df = 3) = 1.25 | 0.74 | 0 | 0 | 1.00 |
| Mean age (years) | 10 | -0.006 (-0.036, 0.025) | 0.72 |  |  |  |
| Body mass index (kg/m<sup>2</sup>) | 10 | 0.018 (-0.091, 0.127) | 0.74 |  |  |  |
| location (Nigeria as ref.) | 10 | -0.470 (-1.370, 0.430) | 0.31 |  |  |  |

Abbreviations:
CI: Confidence interval,
H<sup>2</sup>: Unaccounted variability/ sampling variability
I<sup>2</sup>: Residual heterogeneity / unaccounted variability,
n- Number of studies,
OR-Odds ratio
R<sup>2</sup>: Amount of heterogeneity accounted for.
\*I<sup>2</sup>, and \*p-Value related to the test of moderators.
\* Significant at 0.05

**Table 3:** Meta-regression results for moderating effects of mean age, study location, and BMI on the association between junk food consumption and hypertension in West Africa.

| **Moderator** | **n** | **Coefficient (95% CI)** | ***p* Value** | ***I*<sup>2</sup> (%) \*** | ***R*<sup>2</sup> index (%)** | ***H*<sup>2</sup>** |
| --- | --- | --- | --- | --- | --- | --- |
| **Bivariate meta-regression** |  |  |  |  |  |  |
| Mean age (years) | 6 | 0.001 (-0.031, 0.033) | 0.96 | 2 | 0 | 1.00 |
| Body mass index (kg/m<sup>2</sup>) | 6 | 0.043 (-0.048, 0.134) | 0.35 | 0 | 0 | 1.00 |
| Location (Nigeria as ref.) | 6 | 0.329 (-0.220, 0.877) | 0.24 | 0 | 0 | 1.00 |

| Multivariate meta-regression |  |  |  |  |  |  |
| --- | --- | --- | --- | --- | --- | --- |
| **Test of moderators** | 6 | QM (df = 3) = 0.96 | 0.74 | 0 | 0 | 1.00 |
| Mean age (years) | 6 | -0.039 (-0.116, 0.039) | 0.33 |  |  |  |
| Body mass index (kg/m<sup>2</sup>) | 6 | 0.077 (-0.342, 0.495) | 0.72 |  |  |  |
| location (Nigeria as ref.) | 6 | 0.400 (-1.573, 2.374) | 0.69 |  |  |  |

Abbreviations:
CI: Confidence interval,
H<sup>2</sup>: Unaccounted variability/ sampling variability
I<sup>2</sup>: Residual heterogeneity / unaccounted variability,
n- Number of studies,
OR-Odds ratio,
R<sup>2</sup>: Amount of heterogeneity accounted for.
\*I<sup>2</sup>, and \*p-Value related to the test of moderators.
\* Significant at 0.05

**Table 4:**
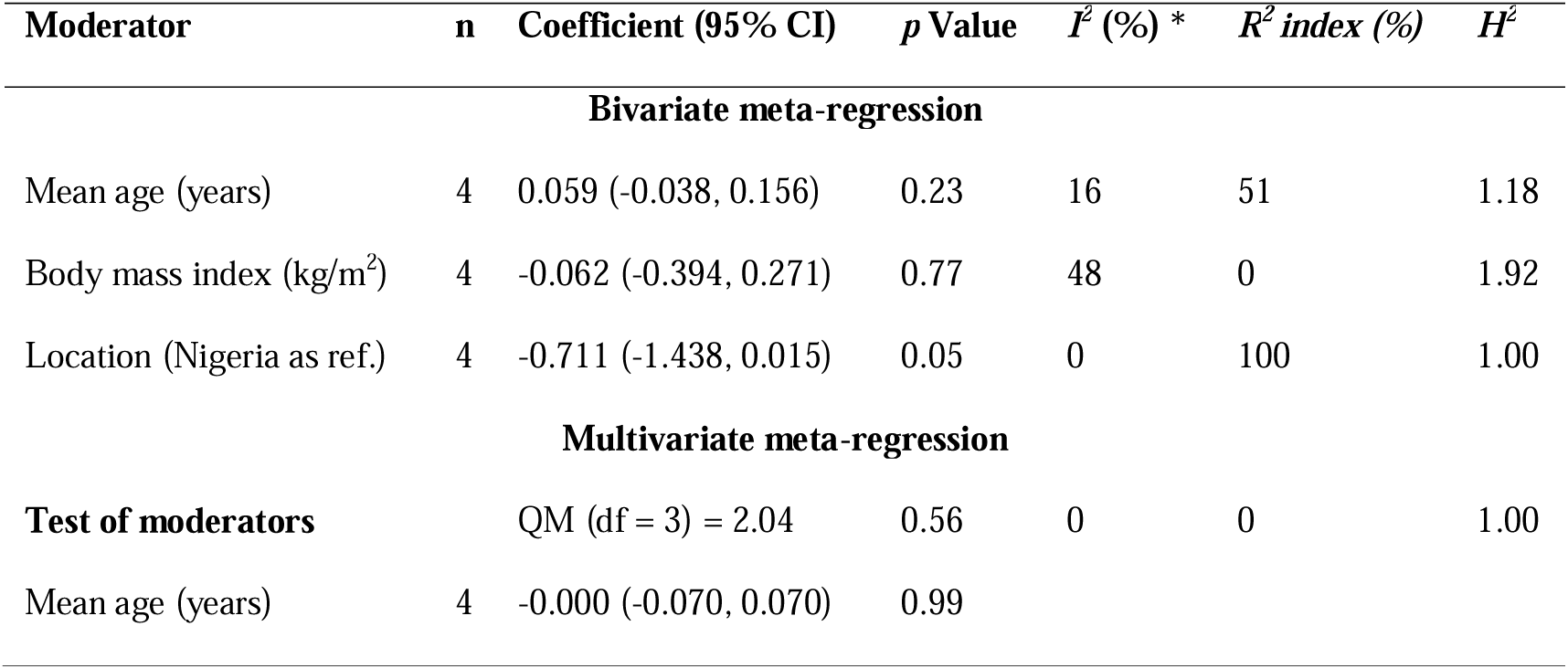
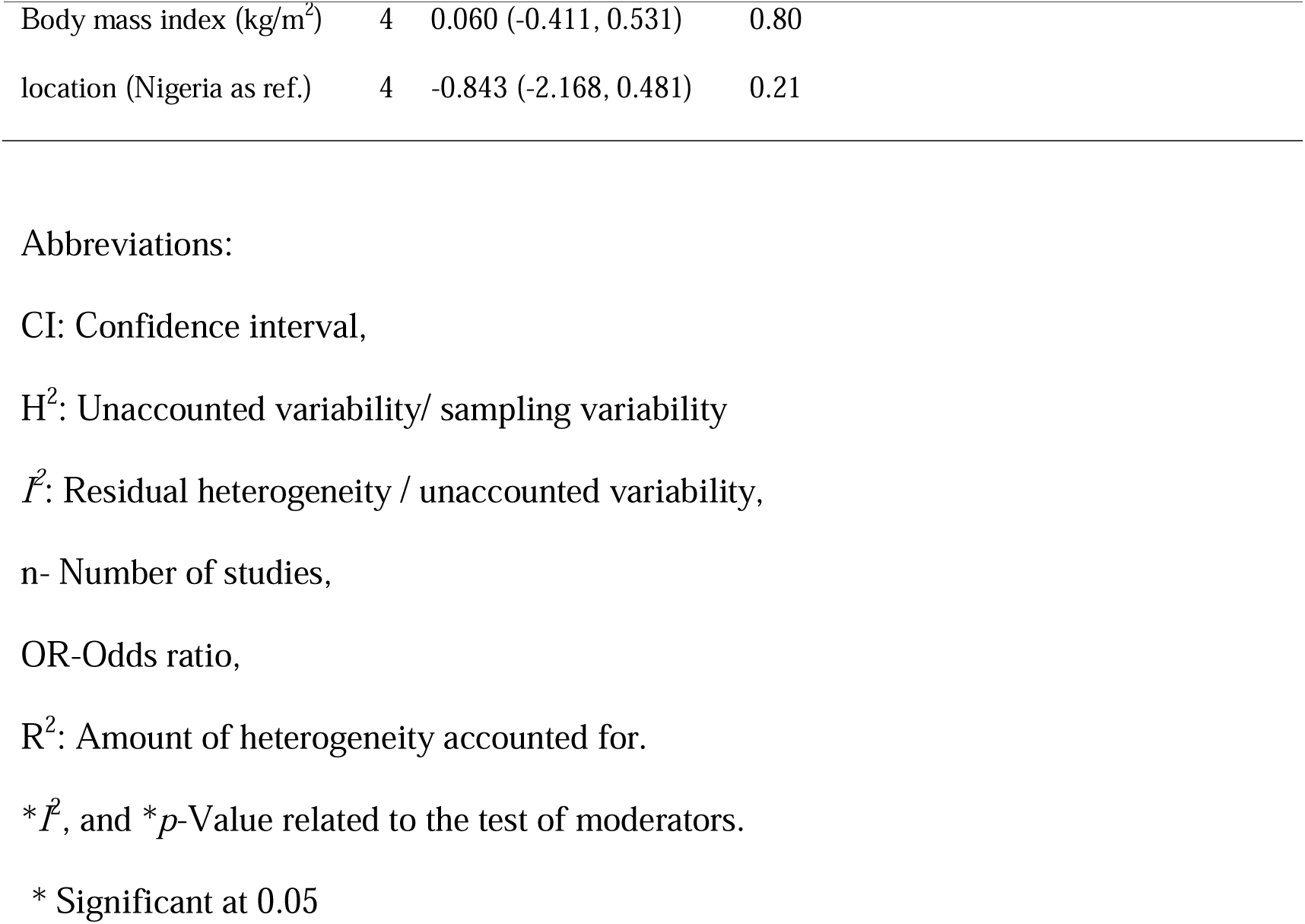
Meta-regression results for moderating effects of mean age, study location, and BMI on the association between red meat consumption and hypertension in West Africa.

| **Moderator** | **n** | **Coefficient (95% CI)** | **p Value** | **I<sup>2</sup> (%) \*** | **R<sup>2</sup> index (%)** | **H<sup>2</sup>** |
| --- | --- | --- | --- | --- | --- | --- |
| **Bivariate meta-regression** |  |  |  |  |  |  |
| Mean age (years) | 4 | 0.059 (-0.038, 0.156) | 0.23 | 16 | 51 | 1.18 |
| Body mass index (kg/m<sup>2</sup>) | 4 | -0.062 (-0.394, 0.271) | 0.77 | 48 | 0 | 1.92 |
| Location (Nigeria as ref.) | 4 | -0.711 (-1.438, 0.015) | 0.05 | 0 | 100 | 1.00 |
| **Multivariate meta-regression** |  |  |  |  |  |  |
| **Test of moderators** |  | QM (df = 3) = 2.04 | 0.56 | 0 | 0 | 1.00 |
| Mean age (years) | 4 | -0.000 (-0.070, 0.070) | 0.99 |  |  |  |

|  |  |  |  |
| --- | --- | --- | --- |
| Body mass index (kg/m<sup>2</sup>) | 4 | 0.060 (-0.411, 0.531) | 0.80 |
| location (Nigeria as ref.) | 4 | -0.843 (-2.168, 0.481) | 0.21 |

Abbreviations:
CI: Confidence interval,
H<sup>2</sup>: Unaccounted variability/ sampling variability
I<sup>2</sup>: Residual heterogeneity / unaccounted variability,
n- Number of studies,
OR-Odds ratio,
R<sup>2</sup>: Amount of heterogeneity accounted for.
\*I<sup>2</sup>, and \*p-Value related to the test of moderators.
\* Significant at 0.05

### Junk food consumption

In the meta-analysis of the association between junk food and the risk of hypertension in West Africa, 6 cross-sectional studies were analyzed with 2,321 participants, of which 781 (33.6%) had hypertension. Junk food was defined as fried food (fried rice, fried chicken, beef, fried yam, and potatoes), fast food, fried snacks (puff-puff, fries, chips), cakes, and burgers. The mean effect sizes across individual studies ranged from 1.28 to 4.95. The meta-analysis demonstrated that consumption of high amounts of junk food was associated with increased odds of hypertension by 41% (OR=1.41; 95% CI: 1.19, 1.67; p < 0.0001, *I^2^*=0%) compared to low consumers in West Africa (**Figure 5**). No heterogeneity or publication bias was observed (**Figure S7**), rank correlation test (p=0.06), or Egger’s regression test (p= 0.26). Subgroup analysis and meta-regression did not suggest a moderating effect by mean population age, BMI, or study location (**Table 4** and **Figures S8 and S9**). This finding indicates that high junk food consumption increases the odds of hypertension by 41% in West Africa.

**Figure 5:**
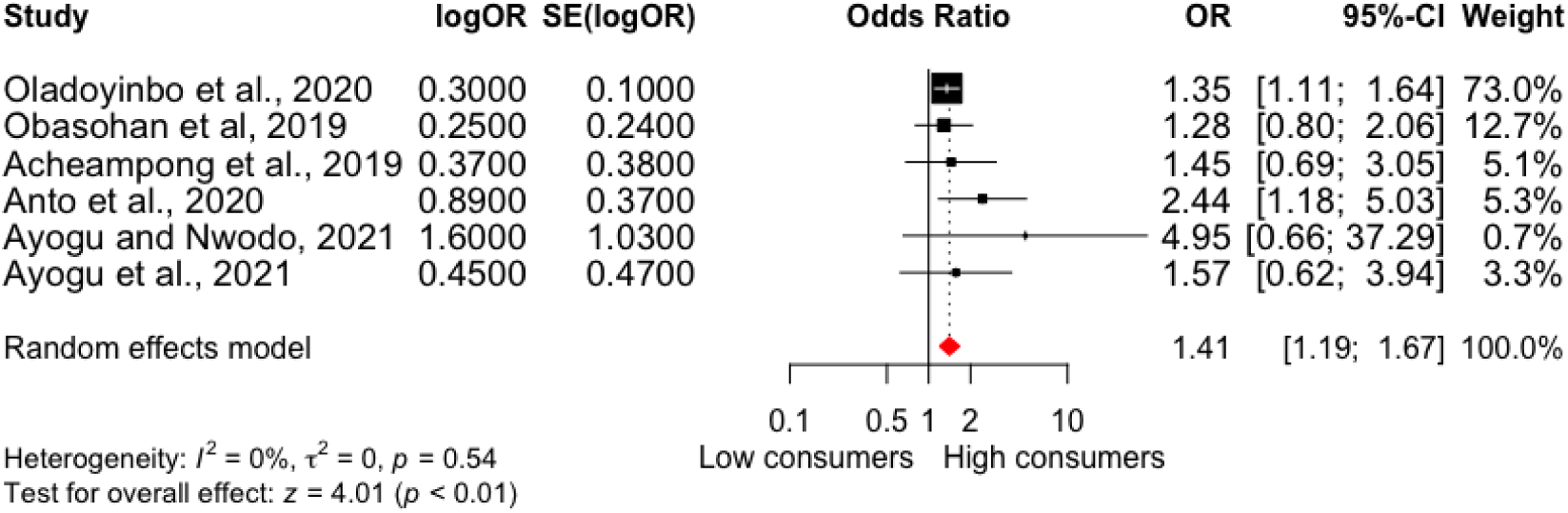
Meta-analysis of the association between junk food consumption and hypertension in West Africa. High junk food consumption increased the odds of hypertension by 41% with no heterogeneity compared to low consumption. CI-Confidence interval, logOR- Treatment effect, OR- Odds ratio, SE(logOR)- Standard error.

### Red meat consumption

The meta-analysis of 4 cross-sectional studies investigating the relationship between red meat consumption and hypertension in West Africa included 10,451 participants, with 1,620 (15.5%) cases of hypertension. Red meat was defined as unprocessed meats from cows, goats, pork, and sheep. The mean effect sizes from the individual studies ranged from 1.06 to 2.56. The meta-analysis suggested that high red meat consumption was associated with a 51% increase in the odds of hypertension (OR=1.51; 95% CI: 1.04, 2.18; p = 0.03, *I^2^*=27%) compared to low consumers with a moderate amount of heterogeneity (**Figure 6**). No heterogeneity or publication bias was observed (**Figure S10**), rank correlation test (p=0.72), or Egger’s regression test (p= 1.00). Subgroup analysis and meta-regression did not suggest moderating effect of mean age, BMI, and study location on the association between the consumption of red meat and hypertension (**Table 5 and Figures S11 and S12**). This highlights that consuming a high amount of red meat significantly increases the odds of hypertension in West Africa by 51%.

**Figure 6:**
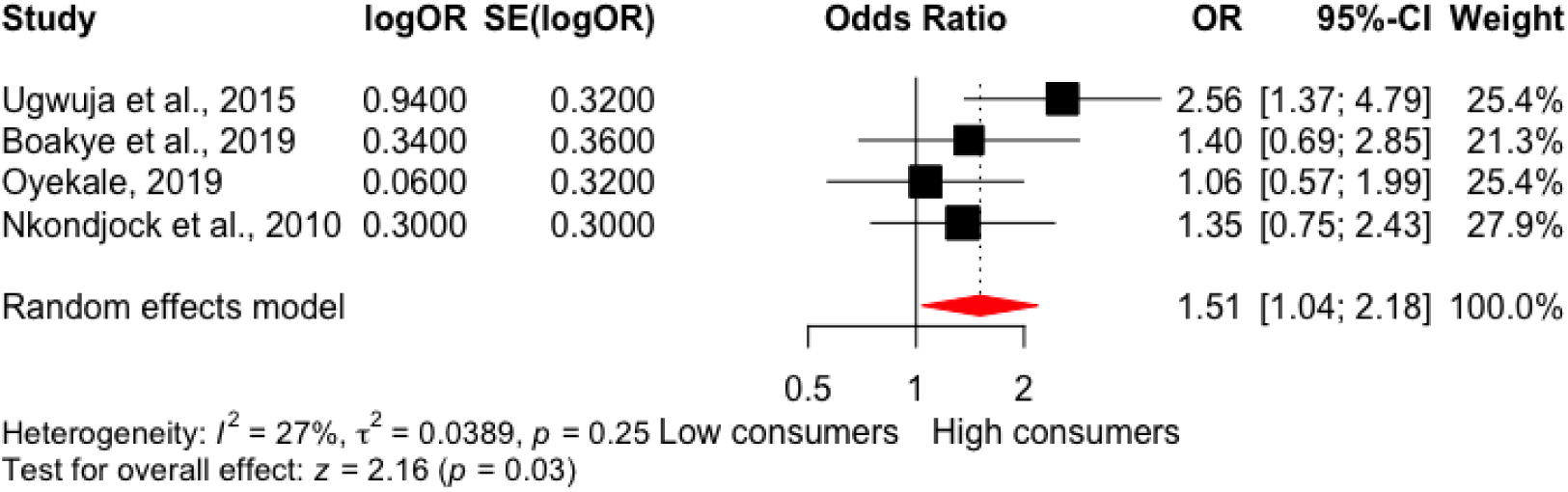
Meta-analysis of the association between red meat consumption and hypertension in West Africa. High consumption of red meat increased the odds of hypertension by 51%, with moderate heterogeneity compared to low consumption. CI-Confidence interval, logOR- Treatment effect, OR- Odds ratio, SE(logOR)- Standard error.

**Table 5:**
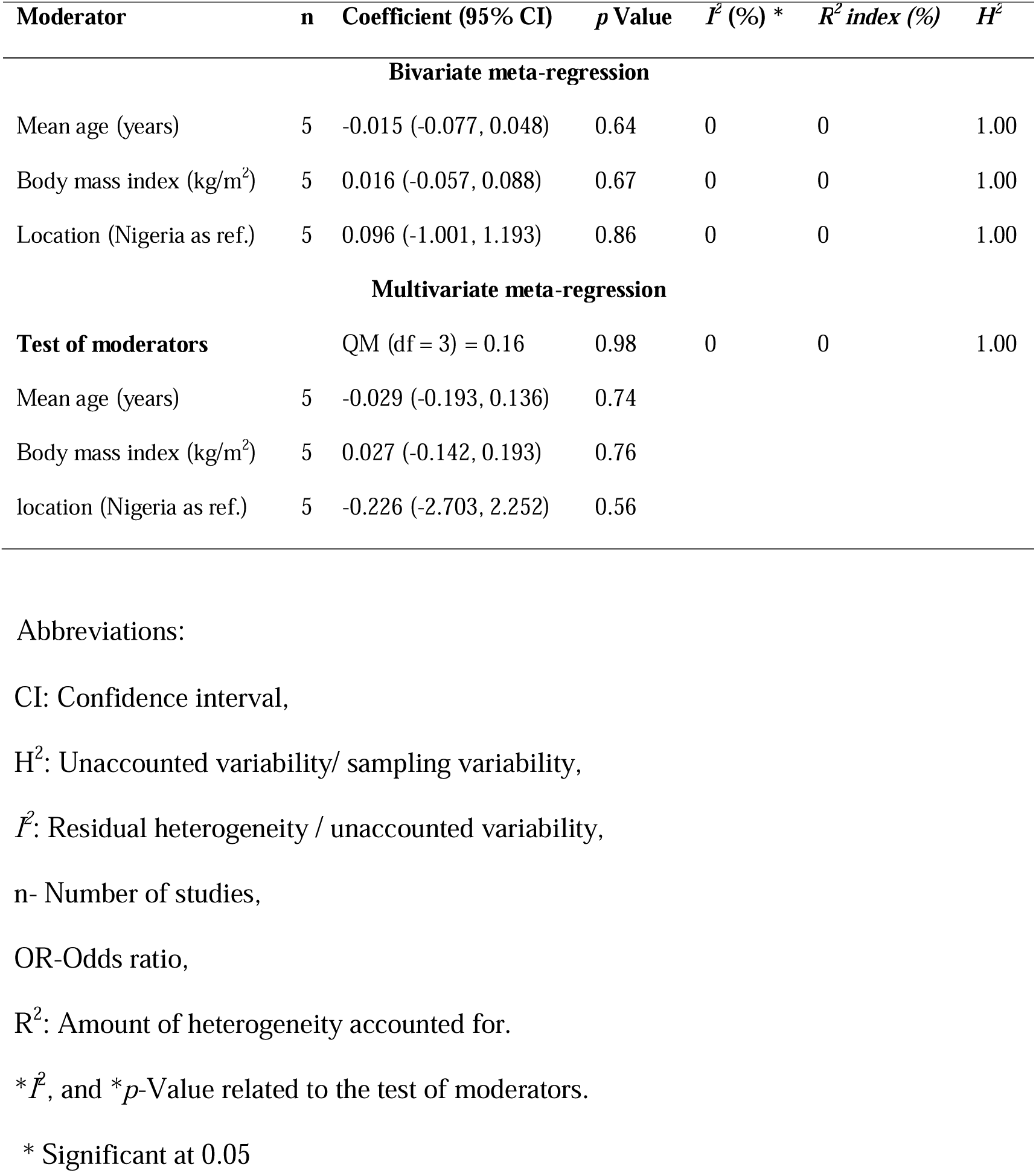
Meta-regression results for moderating effects of mean age, study location, and BMI on the association between dietary fat consumption and hypertension in West Africa.

### Dietary fat consumption

The meta-analysis of 5 cross-sectional studies examining the association between dietary fat and hypertension in West Africa included 5,233 participants, including 1,094 (20.9%) cases of hypertension. Dietary fat was defined as saturated oil, coconut oil, palm and palm oil, butter, lard, margarine, and groundnut oil. Effect sizes from the individual studies ranged from 1.60 to 2.08. The meta-analysis demonstrated that high dietary fat consumption was associated with 76% higher odds of hypertension (OR=1.76; 95% CI: 1.44, 2.14; p < 0.0001, *I*^2^ = 0%) compared to low consumers (**Figure 7**). No heterogeneity or publication bias was demonstrated by the funnel plot (**Figure S13)**, Egger’s regression test (p= 0.89), and Begg’s test (p= 0.48) in the meta-analysis. Subgroup analysis and meta-regression did not suggest a moderating effect on the mean population age, BMI, or study location (**Table 5** and **Figures S14 and S15**). This suggests that high dietary fat consumption increases the odds of hypertension by 76% in West Africa.

**Figure 7:**
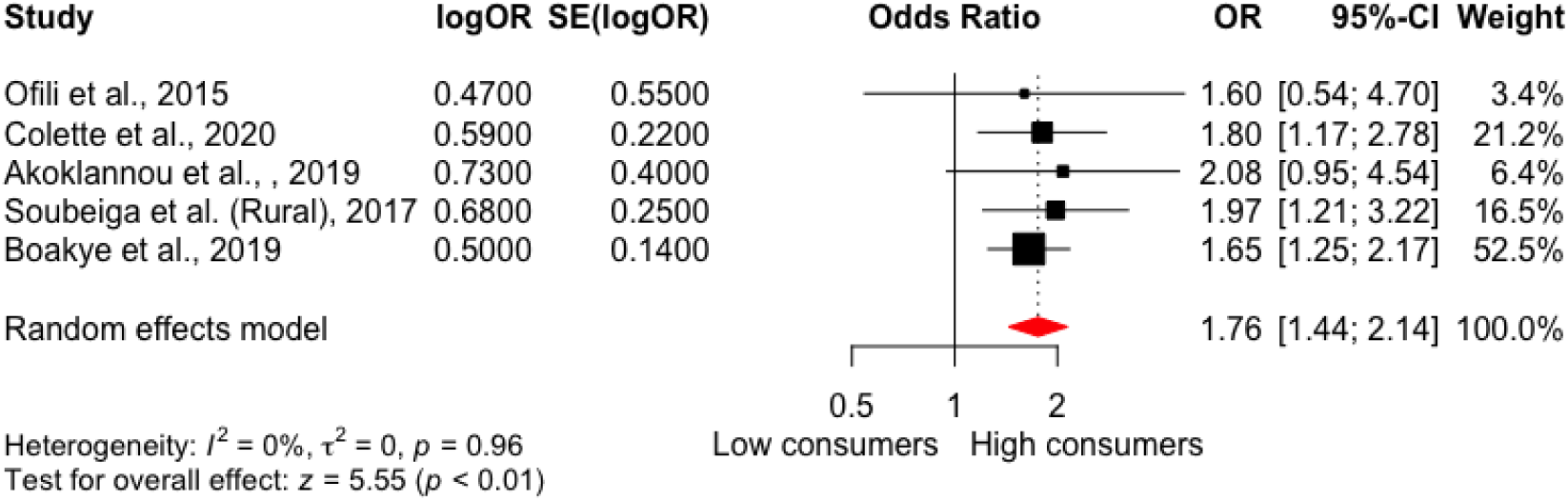
M-analysis of the association between dietary fat consumption and hypertension in West Africa. High dietary fat consumption increased the odds of hypertension by 76% with no heterogeneity compared to low consumption. CI- Confidence interval, logOR- Treatment effect, OR- Odds ratio, SE(logOR)- Standard error.

### Alcohol consumption

A total of 8 cross-sectional studies (n= 6,263 participants, including 2,119 (33.8%) cases of hypertension) reported on the association between alcohol and odds of hypertension were included in the meta-analysis, with effect sizes ranging from 0.82 to 2.03. Compared to low consumers, the meta-analysis demonstrated that high consumption of alcohol increased the odds of hypertension by 17% (OR= 1.17, 95% CI: 1.03, 1.32; p= 0.013, *I*^2^ = 0%) (**Figure 8**). No heterogeneity or publication bias was demonstrated by the funnel plot (**Figure S16)**, Egger’s regression test (p= 0.46), and rank correlation test (p= 0.17) in the meta-analysis. Subgroup analysis and meta-regression did not suggest a moderating effect on the mean population age, BMI, or study location (**Table 6** and **Figures S17 and S18**). This suggests that high alcohol consumption increases the odds of hypertension by 17% in West Africa.

**Figure 8:**
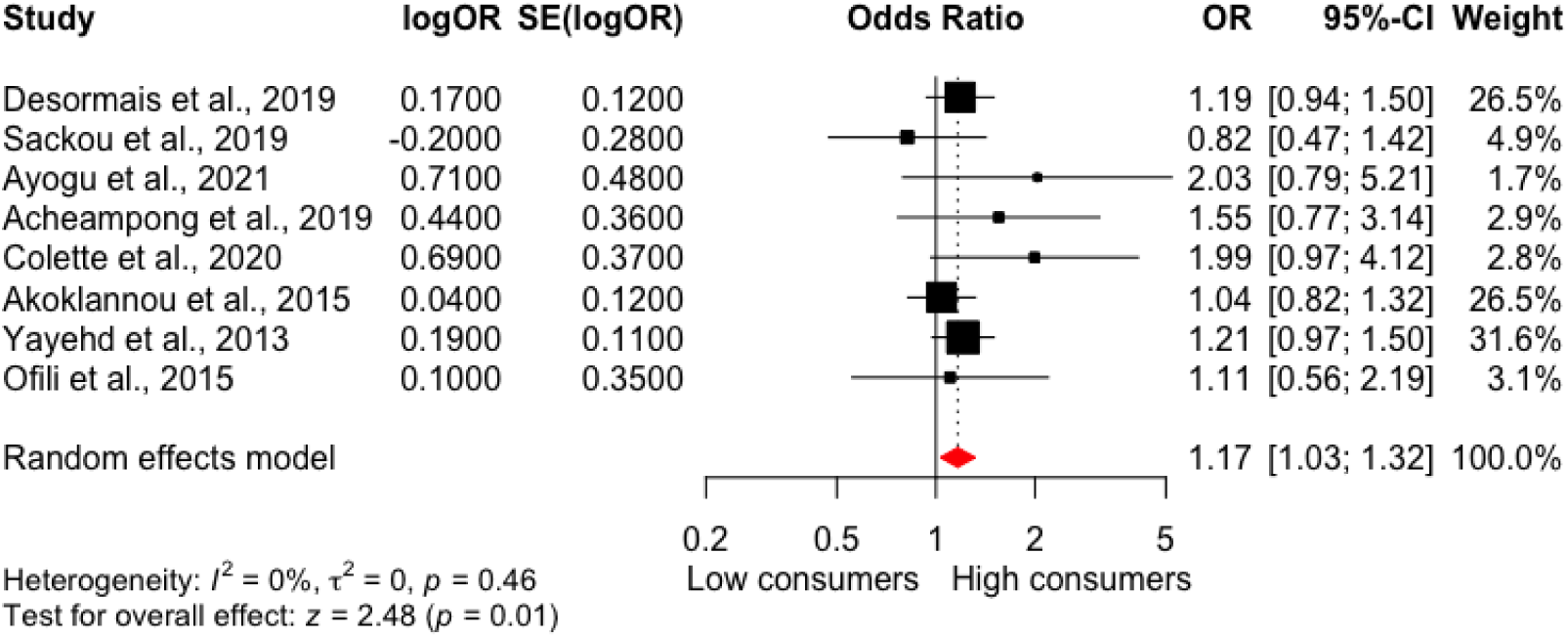
Meta-analysis of the association between alcohol consumption and hypertension in West Africa. High alcohol consumption increased the odds of hypertension by 17% compared to low consumers with no amount of heterogeneity. CI- Confidence interval, logOR- Treatment effect, OR- Odds ratio, SE(logOR)- Standard error.

**Table 6:** Meta-regression results for moderating effects of mean age, study location, and BMI on the association between alcohol consumption and hypertension in West Africa.

| **Moderator** | **n** | **Coefficient (95% CI)** | **p Value** | **$I^2$ (%) \*** | **$R^2$ index (%)** | **$H^2$** |
| --- | --- | --- | --- | --- | --- | --- |
| **Bivariate meta-regression** |  |  |  |  |  |  |
| Mean age (years) | 8 | 0.017 (-1.653, 0.576) | 1.22 | 0 | 0 | 1.00 |
| Body mass index (kg/m<sup>2</sup>) | 8 | 0.000 (-0.050, 0.050) | 0.99 | 10 | 0 | 1.12 |
| Location (Nigeria as ref.) | 8 | -0.167 (-0.741, 0.407) | 0.57 | 6 | 0 | 1.07 |
| **Multivariate meta-regression** |  |  |  |  |  |  |
| **Test of moderators** |  | QM (df = 3) = 0.30 | 0.96 | 0 | 0 | 1.00 |
| Mean age (years) | 8 | 0.012 (-0.053, 0.076) | 0.72 |  |  |  |
| Body mass index (kg/m<sup>2</sup>) | 8 | -0.008 (-0.124, 0.108) | 0.89 |  |  |  |
| location (Nigeria as ref.) | 8 | -0.102 (-1.152, 0.949) | 0.85 |  |  |  |

Abbreviations:
CI: Confidence interval,
$H^2$ : Unaccounted variability/ sampling variability,
$I^2$ : Residual heterogeneity/unaccounted variability,
n- Number of studies,
OR-Odds ratio,
$R^2$ : Amount of heterogeneity accounted for.
$*I^2$ , and $*p$ -Value related to the test of moderators.
\* Significant at 0.05

## Discussion

This systematic review and meta-analysis provide regional-specific evidence to support nutritional guidelines and the development of prevention strategies and tools to reduce the prevalence of hypertension in West Africa. Thirty-one (31) cross-sectional studies with 48,809 participants, including 12,898 (26.4%) adults with hypertension, reported the association between common dietary factors (fruit and vegetables, dietary salt, junk food, red meat, dietary fat, and alcohol) and odds of hypertension. Remarkably, this is the first systematic review and meta-analysis to comprehensively synthesize and analyze the impact of specific dietary factors on hypertension in adults living in West Africa. The results indicate that high consumption of dietary fat, red meat, junk food, dietary salt, and alcohol were associated with an increased likelihood of hypertension in West Africa, while high fruit and vegetable consumption was protective against hypertension. Sensitivity analyses within all dietary factors suggest robust and consistent associations, although some moderators were identified.

In the analysis of fruit and vegetable consumption, the single “healthy” food that was available for analysis, we reported that high fruit and vegetable intake is associated with a reduced risk of hypertension by 20%. This finding is consistent with previous meta-analyses conducted in other regions of the world, such as a meta-analysis of cohort studies by Wu et al. and Schwingshackl et al., which found that high fruit and vegetable intake was associated with a reduction in the incident risk of hypertension, compared to the lowest consumers [36, 37]. According to the meta-analysis of Schwingshackl et al., involving 7 cohort studies for vegetables and fruits, involving 94,772 incident cases of hypertension, a high intake of ‘vegetables and fruits’ reduce the risk of hypertension by ∼7% [37] while the meta-analysis by Wu et al., involving 6 cohort studies demonstrated that high intake of fruit and vegetable reduced the incident of hypertension by 10% [36]. However, both studies reported a moderate to a significant amount of heterogeneity, which may be due to increased diversity in study locations and cultures. In contrast, the observed heterogeneity in effect sizes of our study within a focused geographic region and study population reported no amount of heterogeneity; however, the observed finding that fruits and vegetables have a protective effect against hypertension may be less pronounced in elderly individuals is interesting and warrants further investigation. The results of our meta-analysis agree with the biochemical evidence linking the phytochemicals, vitamins, minerals, and fibers in fruit and vegetables that improved vascular function and antioxidant activity, ultimately reducing the risk of hypertension [38–48].

Conversely, in our analysis of less healthy dietary factors □ many of which are common in processed and convenience foods (i.e., dietary salt, saturated fats, red meat, junk food, and alcohol) □ that have been associated with increased odds of hypertension in studies outside of West Africa, we also report increased odds of hypertension in West African studies. In our meta-analysis of dietary salt, the odds of developing hypertension were increased by 25% in individuals with high dietary salt consumption compared to those with low consumption in West Africa. Our findings agree with previous meta-analyses by Subasinghe et al. and Zhang et al., which reported a positive association between dietary salt intake and hypertension in various populations [49, 50]. The meta-analysis by Subasinghe et al., with 18 studies in rural and urban populations of low to middle-income countries, reported that high dietary salt intake was associated with a 33% increased risk of hypertension compared to the lowest consumers [49], while Zhang et al., reported that among 23 studies in China, high salt intake was associated with an almost 4-fold increase in odds of hypertension [50]. Both studies found significant heterogeneity. In contrast, this meta-analysis found some heterogeneity in effect sizes among the included studies, although this was only moderate. Subgroup analysis and meta-regression suggested that the moderating effect of mean age, BMI, and study location were not significant. The results of our meta-analysis align with the pathophysiology that links salt/sodium intake to hypertension through its effect on renal salt and water retention, elevated plasma sodium levels, and increased salt sensitivity, which can lead to the expansion of extracellular fluid volume, microvascular endothelial inflammation, and structural changes in autonomic and small resistant arteries, resulting in increased systemic peripheral resistance and functional abnormalities of the cardiovascular system and hypertension [51–57].

Similarly, we report that high consumption of junk food is associated with hypertension and increases the odds of hypertension in West African adults by 41%. This finding agrees with biochemical evidence that has linked junk foods, which often contain high amounts of saturated and trans fats, sodium, and by-products of oxidation, which can induce increased blood pressure in different mechanisms [52, 58–63]. In addition, our work agrees with previous work from outside West Africa. A cross-sectional study with 9221 Korean adults in Korea reported that high consumption of fried food led to elevated blood pressure in men and women (odds ratio of 1.62 and 2.20, respectively), compared to infrequent consumers [64], while a meta-analysis of 11 studies (8 cohort and 3 cross-sectional studies) involving 222,544 participants reported a similar positive association that suggests that fried food increased risk of hypertension by 20% compared to low consumers [65]. The absence of heterogeneity in our meta-analysis of the association between junk food and hypertension in West Africa suggests that the findings are robust and not influenced by factors such as study design, population characteristics, or other potential sources of variation. However, it is important to note that the number of studies included in the meta-analysis is relatively small. The findings of our work provide evidence that the association between junk food is associated with an increased risk of hypertension in West Africa.

Our study adds to the growing body of evidence from meta-analyses investigating the association between red meat consumption and hypertension. We found that high red meat consumption was associated with increased odds of hypertension in West Africa. Specifically, our meta-analysis of 4 studies found a 51% increase in the odds of hypertension among high red meat consumers compared to low consumers, which aligns with the meta-analysis conducted by Schwingshackl and Zhang that reported a 15-22% increase in the risk of hypertension in high red meat consumers, compared to low consumers[37], [66]. Schwingshackl et al. conducted a meta-analysis of 7 cohort studies involving 97,745 hypertension cases on food groups and risk of hypertension and reported a positive association between the risk of hypertension and red meat intake by 15%, with substantial heterogeneity (*I^2^*= 84%) [37]. Similarly, Zhang et al. conducted a meta-analysis of 9 cohort studies on red meat, poultry, and egg consumption with the risk of hypertension and reported that high consumption of red meat increased the risk of hypertension by 22%, with substantial heterogeneity (*I^2^* =75%) among the included studies. Our study observed moderate heterogeneity; however, the moderators examined in our meta-analysis did not significantly affect the relationship and effect size. The result of our meta-analysis reflects the proposed biological effect of high meat consumption on hypertension. Meat contains components such as saturated fat, cholesterol, high sodium levels in processed meat, haem iron, and substances formed during cooking or processing of meat that can contribute to the development of hypertension, such as heterocyclic amines, advanced glycosylation end-products, acrylamides, and trimethylamine-N-oxide. These components can affect blood lipids and lead to insulin resistance and inflammation, ultimately increasing the risk of hypertension [67–75]. In short, we confirm that in West Africa, as in other nations, high red meat consumption is associated with an increased likelihood of hypertension in adults.

Additionally, several epidemiological studies have reported an association between dietary fat and the risk of hypertension. Indeed, prospective cohort studies by Yuan et al. and Wang et al. found that high consumption of dietary fat contributed to elevated blood lipids levels and increased the risk of hypertension, with odds ratios of 1.40-1.78, suggesting a greater than 40% increase in hypertension risk associated with high dietary fat consumption [62, 76]. Our meta-analysis of 5 cross-sectional studies supports the translation of this evidence to West Africa, specifically, that high dietary fat consumption may increase the odds of hypertension by 76%. Although our analysis could not differentiate between saturated and unsaturated fats, given the high consumption of saturated fats in West Africa through traditional cooking methods [77–80], that rely on palm and palm kernel oils, groundnut oil, coconut oil, butter, and animal fat, it is likely that saturated fats were the primary form of fat consumed. This aligns with biochemical and pathophysiological evidence that suggests that diets high in saturated fats can lead to increased levels of blood LDL-cholesterol and triglycerides, the development of atheroma in the walls of blood vessels, oxidative stress, and inflammations of the vessels wall, causing stiffness and narrowing of the lumen with reduction of vascular elasticity and lead to increased vascular resistance and hypertension [70, 81–83]. Therefore, our study supports the hypothesis that high dietary fat consumption, particularly saturated fats, is associated with an increased risk of hypertension in West Africa.

Finally, high alcohol consumption increases the odds of hypertension by 17% in West Africa in our meta-analysis. This finding is consistent with previous studies conducted in other regions, which have also reported a positive association between alcohol consumption and hypertension. A meta-analysis of 20 cohort studies (n=361,254 participants and 90,160 incident cases of hypertension) by Roerecke et al. reported that alcohol consumption increases the risk of hypertension by >19% between men and women who drank 1-2 drinks per day in comparison with abstainers with a significant amount of heterogeneity [84]. The absence of heterogeneity or publication bias suggests that the results are reliable and robust, providing strong evidence for the relationship between alcohol consumption and hypertension in West Africa. Subgroup analysis and meta-regression were conducted to explore whether mean population age, BMI, or study location influenced the relationship between alcohol consumption and hypertension. However, no significant moderating effect was found, indicating that the relationship between high alcohol consumption and hypertension is consistent across different population subgroups in West Africa. Our meta-analysis aligns with pathophysiological evidence that alcohol stimulates renin release, which produces angiotensin II, a potent vasoconstrictor, and aldosterone increases sodium retention and water reabsorption. These effects can increase blood pressure and hypertension over time [85, 86]. Also, the production of reactive oxygen species (ROS) and reactive nitrogen species (RNS) during alcohol metabolism can lead to oxidative damage to the endothelium and vascular smooth muscle, resulting in impaired vascular function and increased vascular resistance [87, 88].

This systematic review and meta-analysis provide regional support to the dietary recommendations set forth by the World Health Organisation (WHO) and the 2014 Nigerian National Nutritional Guidelines for the reduction of hypertension. The recommendations include consuming at least 400g or five portions of fruits and vegetables per day, limiting salt intalk to less than 5g per day, reducing consumption of fried and fast foods, restricting red meat consumption to no more than 350-500g or three portions per week and an avoiding processed meat, keeping total dietary fat intake to less than 30% of the total energy intake and limiting the intake of alcohol to ≤ 2 drinks/day for men and ≤1 drink/day for women in the US and less than <14 units in the UK [18, 89–94]. These findings provide region-specific evidence into dietary factors and hypertension risk and identified 6 dietary factors (dietary salt, junk food, dietary fat, red meat, and alcohol) as hypertension-inducing dietary factors and fruit and vegetable as protective against hypertension.

## Strengths and limitations

This systematic review and meta-analysis evaluated the association between dietary factors and hypertension across West Africa has two notable limitations that could not be avoided. First, as with most observational data, nutritional data was collected by participant dietary recall, which is subject to bias. Second, with only cross-sectional studies available, we cannot infer causality. Nonetheless, our study has several notable strengths: (1) this is the first systematic review and meta-analysis that has synthesized and evaluated the association between dietary factors and hypertension across West Africa; (2) despite focussing on a specific region of Africa, a large number of studies and participants were included; (3) a diverse selection of dietary factors were identified that reflected foods items common to healthy and unhealthy patterns; and (4) analyses were critically appraised for risk of bias and power to ensure high-quality.

## Conclusion

This systematic review and meta-analysis demonstrated that high consumption of dietary salt, red meat, junk food, dietary fat, and alcohol are associated with an increased likelihood of hypertension in West Africa. In contrast, high fruit and vegetable consumption appears protective. The results of this study will offer regional-specific evidence for developing clinical nutritional assessment tools for clinicians, patients, and researchers and reduce the ever-increasing trend of hypertension in West Africa.

## Data Availability

All data produced in the present work are contained in the manuscript

## Acknowledgments

The authors sincerely thank the Tertiary Education Trust Fund (TETFund), Nigeria, for funding this study. We also acknowledge the contributions of all the authors and researchers whose work was included in this systematic review and meta-analysis. Finally, we thank all the volunteers in included studies across several countries in West Africa for their dedication and commitment.

## Supplementary material

Table S1: Search strategy used in PubMed database.

Table S2: Characteristics of included studies.

Table S3: Quality assessment and ROB of included studies.

Table S4: PRISMA-P checklist: for dietary factors and hypertension in West Africa.

Figures S1–S18: Subgroup analysis and funnel plots investigating heterogeneity.

