## Supplementary Material for "Dietary factors and hypertension risk in West Africa: A systematic review and meta-analysis of observational studies"

**List of Tables and Figures**

Table S1:	Search strategy used in PubMed database.

Table S2:	Characteristics of the included studies in the meta-analysis investigating the association between dietary factors and odds of hypertension.

Table S3:	Quality assessment of studies included in the meta-analysis using the Newcastle-Ottawa Scale adapted for cross-sectional studies for assessing studies in the meta-analysis of the association between dietary factors and hypertension in West Africa.

Figure S1:	Funnel plots of 19 studies that reported on the association between fruit and vegetable consumption and hypertension in West Africa.

Figure S2:	Forest plot of Subgroup analysis of the moderating effect of (A) mean age, (B) BMI, and study location (C) on the association between fruit and vegetable consumption and hypertension in West Africa.

Figure S3:	Bubble plot 19 studies that reported on the association between fruit and vegetable consumption and hypertension.

Figure S4:	Funnel plots of 10 studies that reported on the association between dietary salt consumption and hypertension in West Africa.

Figure S5:	Forest plot of Subgroup analysis of the moderating effect of (A) mean age, (B) BMI, and study location (C) on the association between dietary salt consumption and hypertension in West Africa.

Figure S6	Bubble plot 10 studies that reported on the association between dietary salt consumption and hypertension.

Figure S7:	Funnel plots of 6 studies that reported on the association between fried and fast-food consumption and hypertension in West Africa.

Figure S8:	Forest plot of Subgroup analysis of the moderating effect of (A) mean age, (B) BMI, and study location (C) on the association between fried and fast-food consumption and hypertension in West Africa.

Figure S9:	Bubble plot 6 studies that reported on the association between fried and fast-food consumption and hypertension.

Figure S10:	Funnel plots of 5 studies that reported on the association between red meat consumption and hypertension in West Africa.

Figure S11:	Forest plot of Subgroup analysis of the moderating effect of (A) mean age, (B) BMI, and study location (C) on the association between red meat consumption and hypertension in West Africa.

Figure S12:	Bubble plot 5 studies that reported on the association between red meat consumption and hypertension. The bubble plot illustrates the relationship of moderating effects of (A) mean age, (B) body mass index (BMI), and (C) study location on the association between red meat consumption and hypertension.

Figure S13:	Funnel plots of 5 studies that reported on the association between dietary fat (fatty food) consumption and hypertension in West Africa.

Figure S14:	Forest plot of Subgroup analysis of the moderating effect of (A) mean age, (B) BMI, and study location (C) on the association between dietary fat (fatty food) consumption and hypertension in West Africa.

Figure S15:	Bubble plot 5 studies that reported on the association between dietary fat (fatty food) consumption and hypertension.

Table S4:	Table S4: PRISMA-P (Preferred Reporting Items for Systematic review and Meta-Analysis Protocols) checklist: for dietary factors and hypertension in West Africa.

**Table S1:** Search strategy used in PubMed database.

| PICO Framework | Concept | Search terms |
| --- | --- | --- |
| P- West African nations | 1 | "Africa, Western"[Mesh] OR “West Africa” OR Benin OR "Burkina Faso" OR "Cape Verde" OR "Cote d'Ivoire" OR “Ivory coast” OR Gambia OR Ghana OR Guinea OR Guinea-Bissau OR Liberia OR Mali OR Mauritania OR Niger OR Nigeria OR Senegal OR Sierra Leone OR Togo OR Cameroon |
| I – High dietary exposure | 2 | (("Diet"[Mesh]) OR "Nutrients"[Mesh]) OR "Food"[Mesh] OR Diet* OR nutrition OR nutrient* OR "food habit*" OR "eating habit*" OR lifestyle OR food* OR "dietary pattern*" OR “fatty food*” OR fat* OR DASH OR “Dietary approaches to stop hypertension” OR “table salt*” OR salt* OR egg* OR vegetables OR “fruit and vegetable” OR fruit* OR “healthy diet*” OR “low-fat dairy product*” OR potassium OR sodium OR meat OR starch OR carbohydrate* OR “dietary supplement*” OR sugar* OR alcohol |
| C- Low dietary exposure |  |  |
| O- Hypertension/ high blood pressure | 3 | "Hypertension"[Mesh] OR hypertension OR "high blood pressure" OR "high systolic blood pressure" OR "high diastolic blood pressure" OR SBP OR DBP OR "elevated blood pressure" OR HTN |

**Table S2.** Characteristics of the included studies in the meta-analysis investigating the association between dietary factors and odds of hypertension

| **S/No** | **The first author,**  **published year** | **Study type** | **Study location** | **Mean age** | **Number of**  **Participants** | **Sex** | | **Exposure** | | **Hypertension** | | **Adjusted covariates** | **Result** |
| --- | --- | --- | --- | --- | --- | --- | --- | --- | --- | --- | --- | --- | --- |
|  |  |  |  |  |  | **Male** | **Female** | **Dietary**  **Assessment tools** | **Estimated dietary factor** | **Method of assessment** | **Number of Cases** |  | **(High vs low intake category) OR (95% CI)** |
| 1. | Anto, Owiredu [1] | CS | Ghana | 44 | 527 | Nil | Nil | Structured questionnaire | Fast food | Measured | 204 | Age, ethnicity, and family history of hypertension | FF (2.43; 1.18-5.02) |
| 2. | Sackou, Tiade [2] | CS | Côte d’Ivoire | 34 | 360 | 123 | 170 | WHO Stepwise questionnaire | Fruit & vegetables, alcohol | measured | 67 | NA | FV (>5) 0.87 (0.22-3.45)  Alcohol (0.82; 0.47-1.41) |
| 3. | Ugwuja, Ezenkwa [3] | CS | Nigeria | 42 | 271 | 86 | 185 | Structured questionnaire | Red meat, fish | measured | 62 | NA | Red meat: fish [27(11): 240(51)]; 2.55(1.11-5.83) |
| 4. | Oguoma, Nwose [4] | CS | Nigeria | 40 | 417 | 145 | 272 | WHO Stepwise questionnaire | Fruit & vegetables | measured | 34 | Age, gender, exposure to second-hand smoke and physical activity status | FV (<5) : (1.06; 1.01–1.13), (>5): 0.94(0.90-0.98) |
| 5. | Olawuyi and Adeoye [5] | CS | Nigeria | 43 | 606 | 326 | 280 | WHO Stepwise questionnaire | Fruit & Vegetables | measured | 201 | Age, gender, exposure to second-hand smoke and physical activity status | FV (low): 0.91 (0.48-1.72), (high): 1.10(0.58-2.08) |
| 6. | Olawuyi and Adeoye [5] | CS | Ghana | 46 | 204 | 80 | 124 | Validated questionnaire | Fruit and vegetables | measured | 100 | Age, alcohol status, overweight, family history of hypertension, unemployment, physical activity | FV (<5): 1.51(0.80-2.84), (>5): 0.66(0.35-1.25) |
| 7. | Acheampong, Nyamari [6] | CS | Ghana | 50 | 216 | NA | NA | WHO Stepwise questionnaire | Extra salt  Fruit & vegetables  Fast food,  alcohol | measured | 73 | age, marital status, education status, employment status, alcohol drinking, extra salt intake, fast foods intake, and body mass index | FV (<5): 3.17 [1.05-9.55], (>5): 0.32(0.11-0.93)  Salt: 1.34 [0.64-2.81]  FF: (No): 0.69[0.33-1.45]- (High): (1.45(0.69-3.05)  Alcohol (1.55; 0.77-3.12) |
| 8. | Obasohan, Okorie [7] | CS | Nigeria | 41 | 476 | NA | NA | Food Frequency Questionnaire (FFQ) | Fast Food | measured | 208 | Age, obesity, fast food, education status, MDA place of work | FF=(>3/week): 1.28(0.80 2.031) |
| 9. | Menyanu, Charlton [8] | CS | Ghana | 58 | 4675 | 1954 | 2799 | WHO Stepwise questionnaire | Extra salt use | Measured | 1444 | sex, age, residence, educational level and hypertension prevalence | Salt: 1.11 (0.952–1.299) |
| 10. | Desormais, Amidou [9] | CS | Benin | 43 | 1777 | 695 | 1082 | WHO Stepwise  survey | Fruits and vegetables  Salt intake, alcohol | Measured | 584 | age, sex, country, area, marital status, and previous occupation, tobacco use, BMI, physical activity, diabetes, salt consumption, alcohol consumption, fruits & vegetables | FV (>5): 0.97(0.69-1.36)  Salt (1.23 (1.00–1.50)  Alcohol (1.19; 0.94-1.53) |
| 11. | Simo, Agbor [10] | CS | Cameroon | 53 | 526 | 173 | 353 | WHO step wise approach to surveillance | Fruit& vegetable | Measured | 215 | Age, marital status, gender, education, family history hypertension, smoking status, alcohol consumption, physical activity, BMI, fruit and vegetable consumption | FV (>5): 0.95(0.89-1.03), |
| 12. | Ofili, Ncama [11] | CS | Nigeria | 53 | 134 | 48 | 86 | Questionnaire | Salt, fat, alcohol | Measured | 59 | Age, marital status, gender, education, family history hypertension, smoking status, alcohol consumption, physical activity, BMI, fruit and vegetable consumption, salt | Salt: 1.59 (0.79, 3.20)  Fat: 1.60 (0.54, 4.75)  Alcohol (1.10; 0.55-2.20) |
| 13. | Nkondjock and Bizome [12] | CS | Cameroon | 37 | 571 | 473 | 98 | food frequency questionnaire | Fruit & vegetable  Meat (bush meat, red meat) | Measured | 223 | age, body mass index, rank, vigorous physical activity and total energy intake, the fruit and vegetable | FV (high): 0.82(0.20-2.19)  Meat: 1.35 (0.74–2.47) |
| 14. | Pancha Mbouemboue, Derew [13] | CS | Cameroon | 36 | 700 | 340 | 360 | Semi-structured questionnaire | Salt | Measured | 143 | Age, marital status, gender, education, family history hypertension, smoking status, alcohol consumption, physical activity, BMI, salt, hyperlipidaemia | Salt (high): 1.05 [0.52–2.07] |
| 15. | Charity E. C, Osaretin A.T. E [14] | CS | Nigeria | 39 | 410 | 230 | 180 | WHO STEPS questionnaire | Fruit, vegetable, salt intake | Measured | 205 | Physical activity, fruit and vegetable, current smokers, current alcohol drinkers, salt intake, obesity, diabetes, hypercholesterolemia | FV (>5): 0.15 (0.09, 0.27)  Salt  Never: 0.400 (0.198–0.81)  Often: 2.5 (1.23-5.05) |
| 16. | Oladoyinbo, Abiodun [15] | CS | Nigeria | 35 | 300 | 192 | 108 | structured questionnaire | Snack, puff puff,  fried foods | measured | 144 | BMI, age, smoking, meal skipping, snacking, physical activity, alcohol, | FAST: 1.35 (0.29, 1.65) |
| 17. | Ayogu, Ezeh [16] | CS | Nigeria | 56 | 517 | 213 | 287 | Interviewer administered questionnaire | Fruit, vegetable, nut, legume, alcohol | Measured | 195 | Age, sex, educational level, self-percieved health status, obesity, alcohol intake, fruit, vegetable, nuts , legume | F (>5): 0.64 (0.32–1.27)  V (>5): 0.82 (0.54–1.24)  FV (>5): 0.77(0.54-1.09)  Alcohol (2.04; 0.80-5.20) |
| 18. | Ayogu and Nwodo [17] | CS | Nigeria | 20 | 401 | 188 | 213 | Interviewer method of questionnaire | Snacks, baked food products, fruits, vegetables | Measured | 76 | Age, sex, marital status, Socioeconomic variables of interest were occupation and monthly income, smoking, alcohol consumption, sleep duration and exercise, Meal skipping, consumption of snacks, fruits, vegetables and sugar- containing drinks | Snacks= 1.30 (0.08,21.98)  FF= 4.95 (0.66, 37.13)  FE= 3.15 (0.61, 16.26)  Sugar drinks= 1.57(0.62, 3.93)  Fruit=0.53(0.24, 1.17) |
| 19. | Akoklannou A. H, Yanogo [18] | CS | Benin | 35 | 717 | 327 | 390 | WHO Stepwise survey | Salt  Fatty food,  Fruits and vegetables, alcohol | Measured | 238 | Sex, age, ethinicity, marital status, salt intake, fatty food consumption, physical activity, fruit and vegetable, alcohol consumption, tobacco consumption, BMI, Glucose level | High salt (1.54; 1.07 - 2.21)  Fruit (<5): 1.34(0.66-2.70), (>5): 0.75(0.37-1.52)  fat (2.07; 0.94 - 4.54)  Alcohol (1.04; 0.83-1.30) |
| 20. | Soubeiga, Millogo [19] | CS | Burkina Faso  (Rural) | 44 | 3600 | 1773 | 1827 | WHO Stepwise  approach to Surveillance survey | Butter, lard, margarine, vegetable oil | Measured | 553 | Age, education, marital status, sex, family history of HBP, smoking, physical activity, fats, BMI, HDL cholesterol | High fats: 1.42 (0.89–2.27)  1.98(1.22–3.22) |
| 21. | Yayehd, Damorou [20] |  | Togo | 49 | 2002 | 907 | 1095 | Interviewer administered questionnaire | Salt, alcohol | measured | 734 | Alcohol, Tobacco, salt, obesity, cola, oestrogens | High Salt: 1.4; 1.13–1.72)  Alcohol (1.21; 0.97-1.51) |
| 22. | Colette A., Charles S. J. [21] |  | Benin | 40 | 540 | NA | NA | questionnaire | Salt, fat  Fruit & vegetable, alcohol | Measured | 154 | Place of residence, salt, fruit and vegetables, tobacco, physical use, physical activity, obesity, alcohol intake | F (>5): 0.83 [0.56; 1.23]  Salt (3.40; 1.76-6.57)  Fat (1.81; 1.17-2.81)  Alcohol (2.00; 0.97-4.12) |
| 23. | Shokunbi and Ukangwa [22] | CS | Nigeria | 20 | 488 | 124 | 219 | 24-hour  dietary recall | Puff-puff, eggs, fruits, vegetables, Beef | Measured | 145 | NA | Red meat [ NO- 343(105); YES- 81(64)] OR=2.87(1.92-4.28)  Fried: N0-343(202) vs Yes-94(57): OR-1.08(0.68-1.71)  FV: No-343(275), Yes-94(80)  (Low): 1.41(0.76-2.65), (High)- 0.71(0.38-1.32) |
| 24. | Kingsley Boakye, Comfort Arthur [23] | CS | Ghana | 44 | 242 | 104 | 138 | Structured questionnaires | Salt, fruit & vegetable, palm oil, protein (animal, fish and plant) | Measured | 90 | NA | Salt (High): 1.53(1.05, 2.22)  Fruit: (high)= 0.66(0.52, 0.85)  Veg: (high)= 0.83 (0.64, 1.08)  Red meat=1.40 (0.69, 2.83)  Fat: Palm oil=1.65(1.25, 2.19) |
| 25. | Owiredu, Dontoh [24] | CS | Ghana | 46 | 204 | 80 | 124 | Elaborate pilot-tested questionnaire | Fruit and vegetable | Measured | 100 | Age, sex, highest educational level, marital status, employment status, physical activity, fruits and vegetable intake, alcohol consumption, and family history of hypertension. | FV (low)= 1.51(0.80-2.84)  FV(inverse)= 0.66(0.35, 1.25) |
| 26. | Dorgbetor, Dickson [25] | Survey | Ghana | 36 | 5662 | NA | 5662 | Structured questionnaire | Salted fish | Measured | 918 | Marital status, age, level of education, wealth index, parity, occupation | Salt (Yes)=1.06 [0.90–1.24] |
| 27. | Diendere, Kabore [26] | Survey | Burkina Faso | 44 | 4187 | 2257 | 1930 | WHO STEPS survey tool | Fruit and vegetable | Measured | 774 | living environment, sex, age, marital status, education level, BMI, Physical activity, alcohol use and occupation. | FV (low) = 2.8 (1.2–6.7)  FV (inverse)=0.36[0.15, 0.83) |
| 28. | Modey Amoah, Esinam Okai [27] | CS | Ghana | 62 | 360 | 105 | 255 | Structured questionnaire | Meat, fats, fruit, and vegetables | Measured | 360 | Sex, age, education, knowledge, presence of comorbidity, alcohol, and number of pills taken. |  |
| 29. | Akpa, Okekunle [28] | Survey | Nigeria | 55.4 | 3215 | 1530 | 1685 | Semi-quantitative food frequency questionnaire (FFQ), | Vegetables | Measured | 1727 | Sex, age, education, family history of CVD, ever smoked, ever used alcohol, physical inactivity, BMI, Diabetes, Dyslipidemia | V(high)= 0.96(0.59, 1.57) |
| 30. | Akpa, Okekunle [28] | Survey | Ghana | 53.7 | 3214 | 1541 | 1673 | Semi-quantitative food frequency questionnaire (FFQ), | Vegetables | Measured | 1240 | Sex, age, education, family history of CVD, ever smoked, ever used alcohol, physical inactivity, BMI, Diabetes, Dyslipidemia | V(high)= 0.97(0.77, 1.23) |
| 31. | Akpa, Okekunle [28] | Survey | Burkina Faso | 49.8 | 2097 | 1056 | 1041 | Semi-quantitative food frequency questionnaire (FFQ), | Vegetables | Measured | 345 | Sex, age, education, family history of CVD, ever smoked, ever used alcohol, physical inactivity, BMI, Diabetes, Dyslipidemia | V(high)= 1.09(0.13, 9.19) |
| 32. | Makinde and Babalola [29] | CS | Nigeria |  | 397 | 108 | 289 | Food frequency questionnaire | Meat | Measured | 39 | NA | 0.87 (0.37, 1.96) |
| 33. | Oyekale [30] | Survey | Ghana |  | 9367 | NA | 9367 | Food frequency questionnaire | Salted meat | Measured | 1244 | Age, BMI | β= 0.06, z= 2.03,  OR= 1.06 (1.00, 1.12) |
|  | **Total** |  |  |  | **48,809** |  |  |  |  |  | **12,898** |  |  |

Abbreviations: CS= Cross-sectional study, FF= Fried/fast food, FV= Fruit and vegetable, V= vegetable, $\beta$= Beta-coefficient, z=Z-score

**Table S3:** Quality assessment of studies included in the meta-analysis using the Newcastle-Ottawa Scale adapted for cross-sectional studies for assessing studies in the meta-analysis of the association between dietary factors and hypertension in West Africa.

| **Study** | **Selection of study groups** | | | | **Comparability** | **Outcome** | | **Total score** | **Study**  **Quality**  **(*9)** | **Risk of Bias** |
| --- | --- | --- | --- | --- | --- | --- | --- | --- | --- | --- |
|  | **Representativeness of the sample**  **(*)** | **Sample size**  **(*)** | **Non-Response rate**  **(*)** | **Ascertainment of the screening/surveillance tool**  **(**)** | **Potential**  **Confounders**  **(**)** | **Assessment of**  **Outcome**  **(*)** | **Statistical test**  **(*)** |  |  |  |
| Anto, Owiredu [1] | * | * | * | ** | ** | * | * | ********* (9) |  | Low risk |
| Sackou, Tiade [2] | * | * | * | ** | ** | * | * | ********* (9) |  | Low risk |
| Ugwuja, Ezenkwa [3] | * | * | * | ** | - | * | * | ******* (7) |  | Low risk |
| Oguoma, Nwose [4] | * | * | * | ** | - | * | * | ********* (9) |  | Low risk |
| Olawuyi and Adeoye [5] | * | * | * | ** | ** | * | * | ********* (9) |  | Low risk |
| Olawuyi and Adeoye [5] | * | * | * | ** | ** | * | * | ********* (9) |  | Low risk |
| Acheampong, Nyamari [6] | * | * | * | ** | ** | * | * | ********* (9) |  | Low risk |
| Obasohan, Okorie [7] | * | * | * | ** | ** | * | * | ********* (9) |  | Low risk |
| Menyanu, Charlton [8] | * | * | * | ** | ** | * | * | ********* (9) |  | Low risk |
| Desormais, Amidou [9] | * | * | * | ** | ** | * | * | ********* (9) |  | Low risk |
| Simo, Agbor [10] | * | * | * | ** | ** | * | * | ********* (9) |  | Low risk |
| Ofili, Ncama [11] | * | * | * | ** | ** | * | * | ********* (9) |  | Low risk |
| Nkondjock and Bizome [12] | * | * | * | ** | ** | * | * | ********* (9) |  | Low risk |
| Pancha Mbouemboue, Derew [13] | * | * | * | ** | ** | * | * | ********* (9) |  | Low risk |
| Charity E. C, Osaretin A.T. E [14] | * | * | * | ** | ** | * | * | ********* (9) |  | Low risk |
| Oladoyinbo, Abiodun [15] | * | * | * | ** | ** | * | * | ********* (9) |  | Low risk |
| Ayogu, Ezeh [16] | * | * | * | ** | ** | * | * | ********* (9) |  | Low risk |
| Akoklannou A. H, Yanogo [18] | * | * | * | ** | ** | * | * | ********* (9) |  | Low risk |
| Soubeiga, Millogo [19] | * | * | * | ** | ** | * | * | ********* (9) |  | Low risk |
| Soubeiga, Millogo [19] | * | * | * | ** | ** | * | * | ********* (9) |  | Low risk |
| Yayehd, Damorou [20] | * | * | * | ** | ** | * | * | ********* (9) |  | Low risk |
| Colette A., Charles S. J. [21] | * | * | * | ** | ** | * | * | ********* (9) |  | Low risk |
| Shokunbi and Ukangwa [22] | * | * | * | ** |  | * | * | ******* (7) |  | Low risk |
| Kingsley Boakye, Comfort Arthur [23] | * | * | * | ** |  | * | * | ******* (7) |  | Low risk |
| Owiredu, Dontoh [24] | * | * | * | ** | ** | * | * | ********* (9) |  | Low risk |
| Dorgbetor, Dickson [25] | * | * | * | ** | ** | * | * | ********* (9) |  | Low risk |
| Diendere, Kabore [26] | * | * | * | ** | ** | * | * | ********* (9) |  | Low risk |
| Modey Amoah, Esinam Okai [27] | * | * | * | ** | ** | * | * | ********* (9) |  | Low risk |
| Akpa, Okekunle [28] | * | * | * | ** | ** | * | * | ********* (9) |  | Low risk |
| Akpa, Okekunle [28] | * | * | * | ** | ** | * | * | ********* (9) |  | Low risk |
| Akpa, Okekunle [28] | * | * | * | ** | ** | * | * | ********* (9) |  | Low risk |
| Makinde and Babalola [29] | * | * | * | ** |  | * | * | ******* (7) |  | Low risk |
| Oyekale [30] | * | * | * | ** | ** | * | * | ********* (9) |  | Low risk |

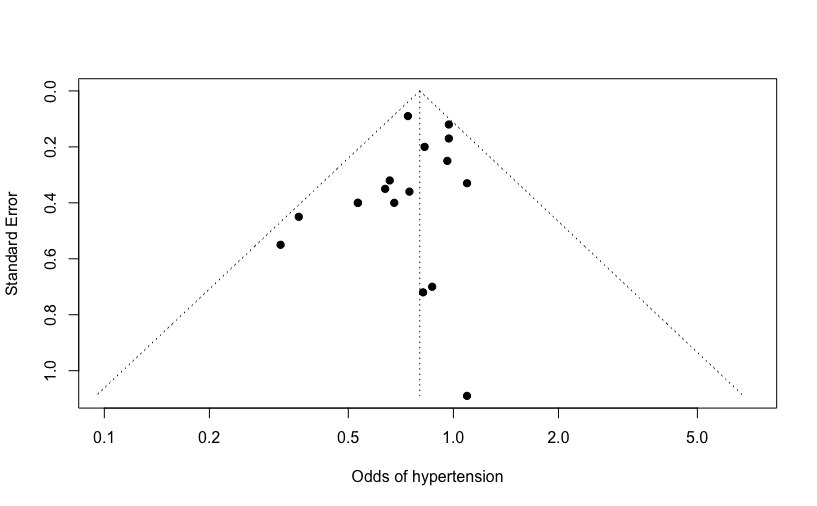

**Figure S1:** Funnel plot of 17 studies reported the association between fruit and vegetable consumption and hypertension in West Africa with an odds ratio (x-axis) vs standard error (y-axis). No publication bias was demonstrated by the funnel plot, Egger’s regression test (p= 0.18), and rank correlation test (p= 0.17) in the meta-analysis.

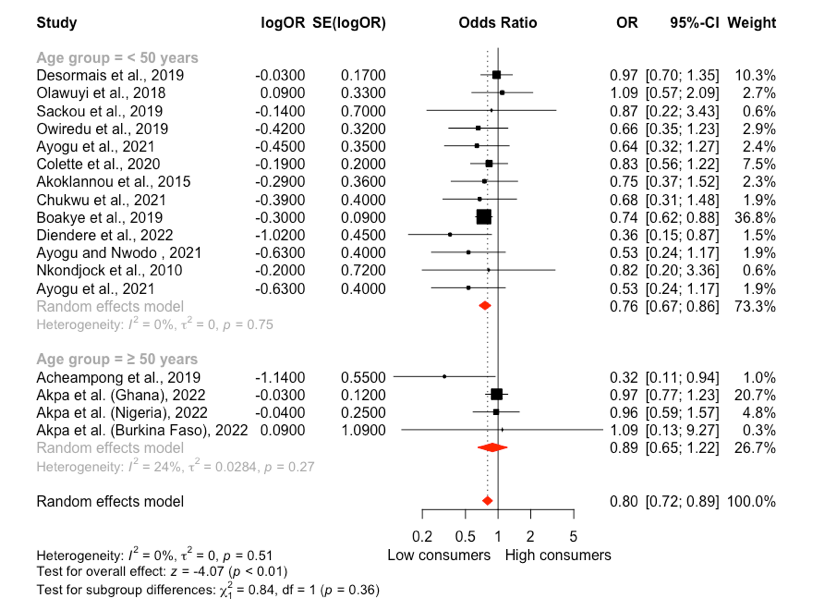

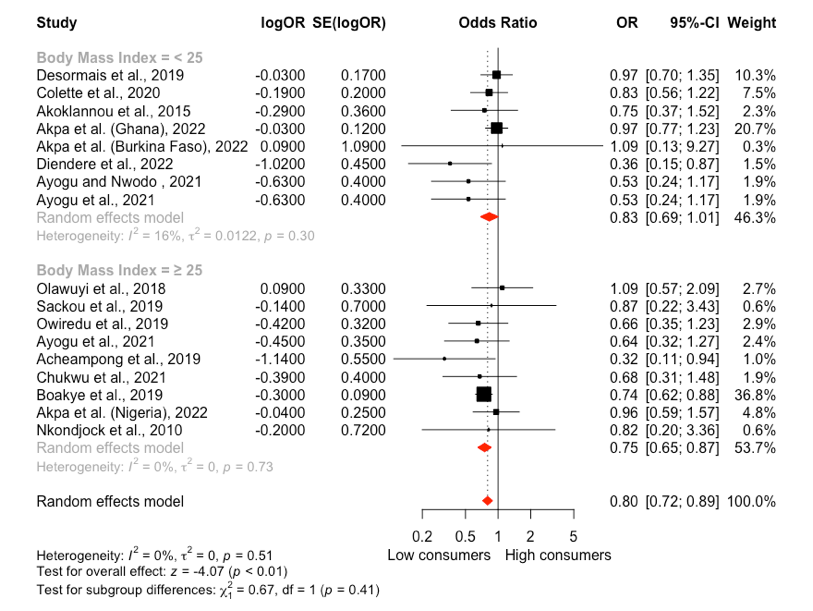

(A) **(B)**

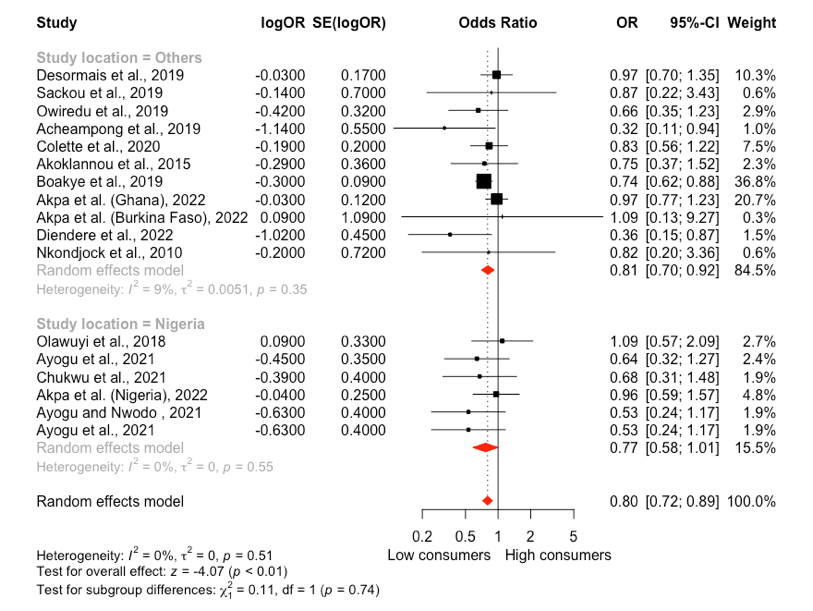

**(C)**

**Figure S2.** Forest plot of Subgroup analysis of the moderating effect of (A) mean age, (B) BMI, and study location (C) on the association between fruit and vegetable consumption and hypertension in West Africa. The result in (A) suggests that mean age significantly moderates the effect size and shows a stronger association in individuals < 50 years than those ≥50 years old. No significant moderating effect was observed for BMI and study location in (B) and (C), respectively. CI: Confidence interval, logOR: Treatment effect, OR: Odds ratio, SE (logOR): Standard error.

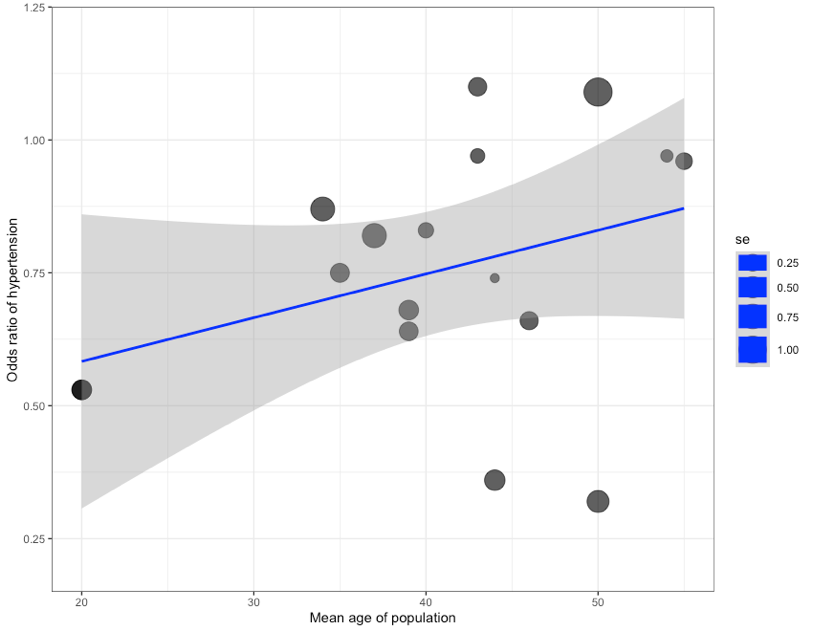

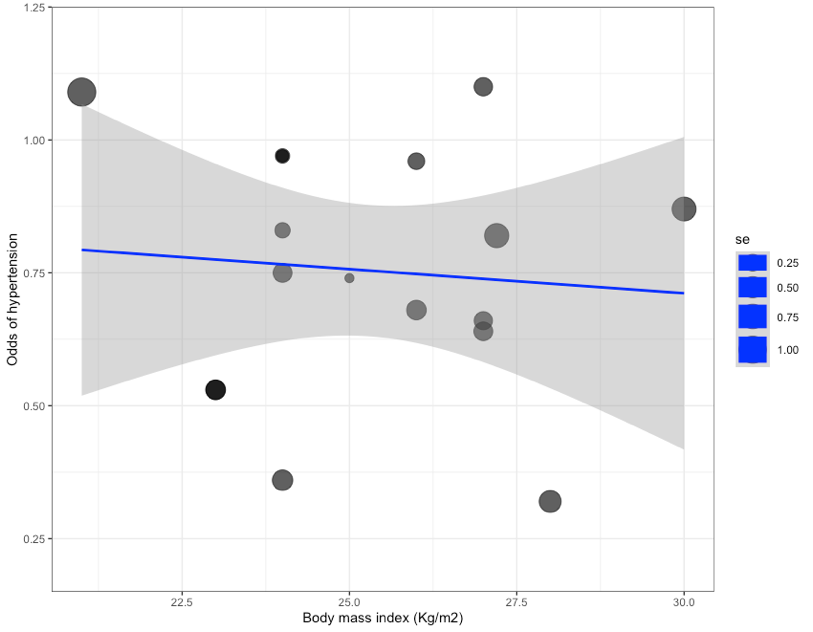

(A) **(B)**

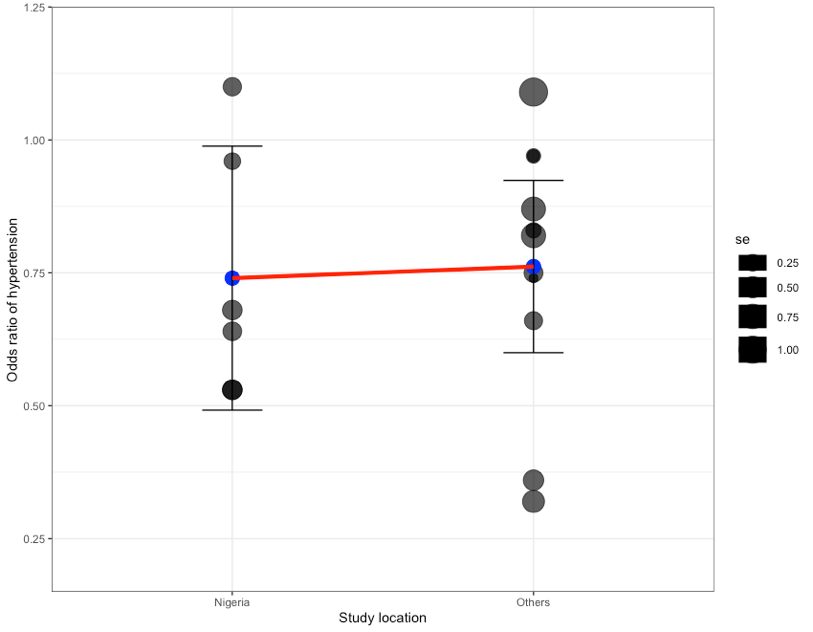

**(C)**

**Figure S3**. Bubble plot of 17 studies reported the association between fruit and vegetable consumption and hypertension. The bubble plot illustrates the relationship of moderating effects of (A) mean age, (B) body mass index (BMI), and (C) study location on the association between fruit & vegetable consumption and hypertension. The bubble plot in (A), (B), and (C) suggests that there is no significant moderating effect of the mean age, BMI, and study location on the relationship between fruit consumption and hypertension.

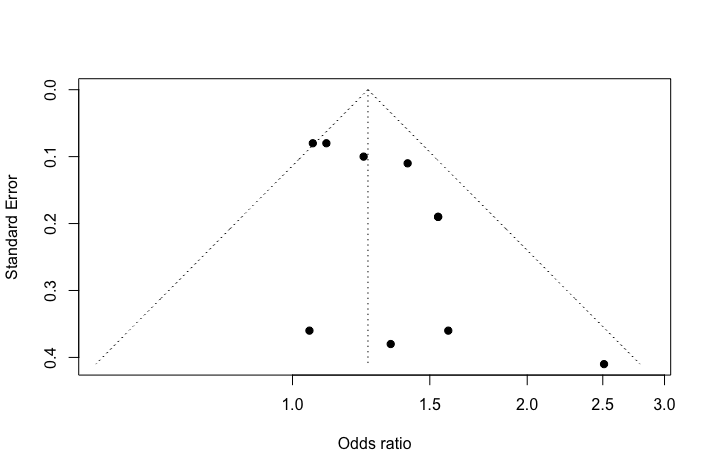

**Figure S4.** Funnel plot of 10 studies reported the association between dietary salt consumption and hypertension in West Africa with an odds ratio (x-axis) vs standard error (y-axis). No publication bias was demonstrated by the funnel plot, Egger’s regression test (p= 0.31), and rank correlation test (p= 0.05) in the meta-analysis.

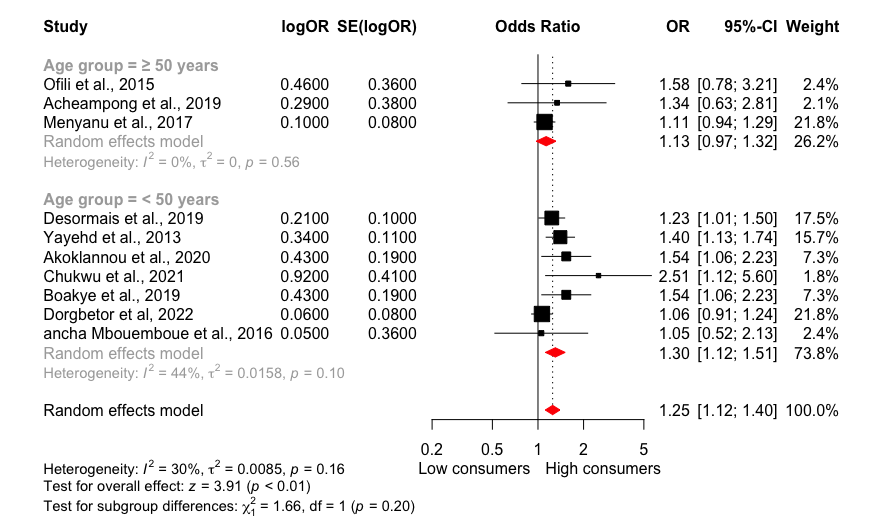

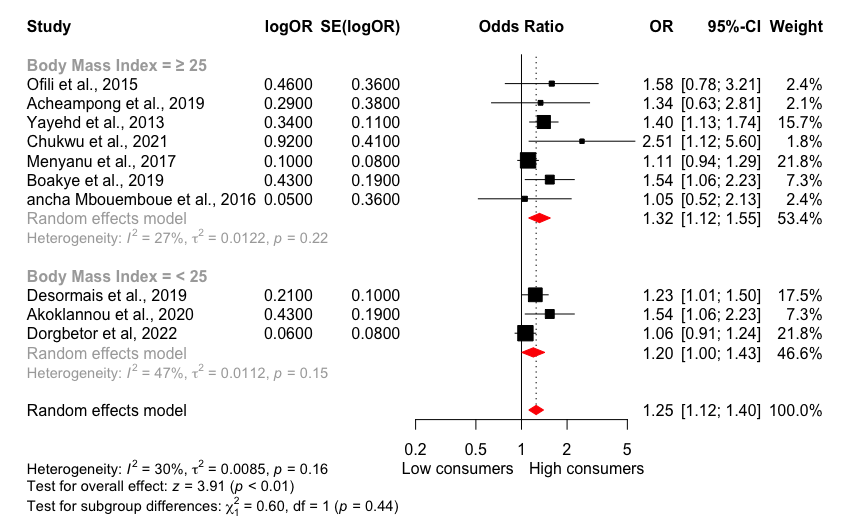

**(A)	(B)**

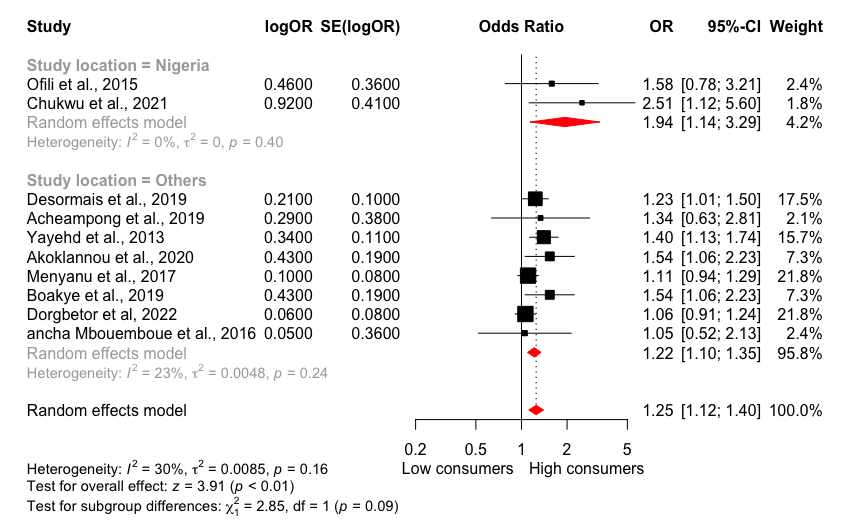

**(C)**

**Figure S5.** Forest plot of Subgroup analysis of the moderating effect of (A) mean age, (B) BMI, and study location (C) on the association between dietary salt consumption and hypertension in West Africa. No significant moderating effect was observed for mean age, BMI, and study location in (A), (B), and (C) on the effect size, respectively. CI: Confidence interval, logOR: Treatment effect, OR: Odds ratio, SE (logOR): Standard error.

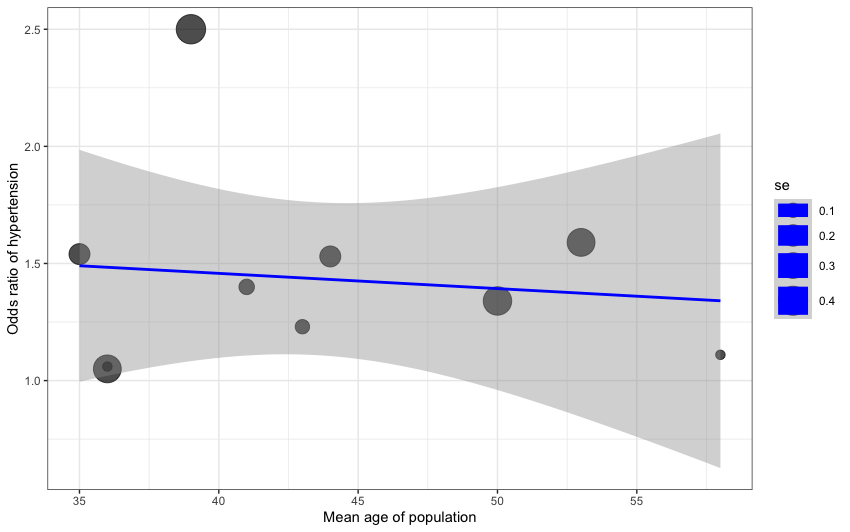

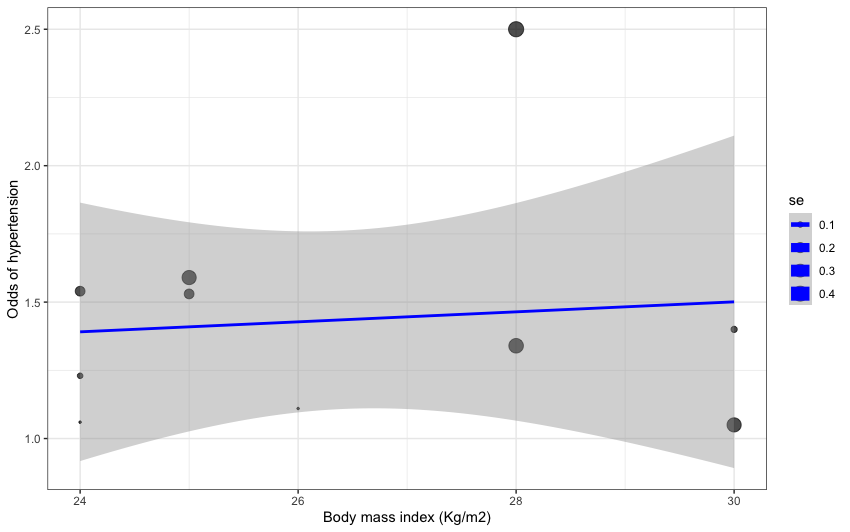

(A) **(B)**

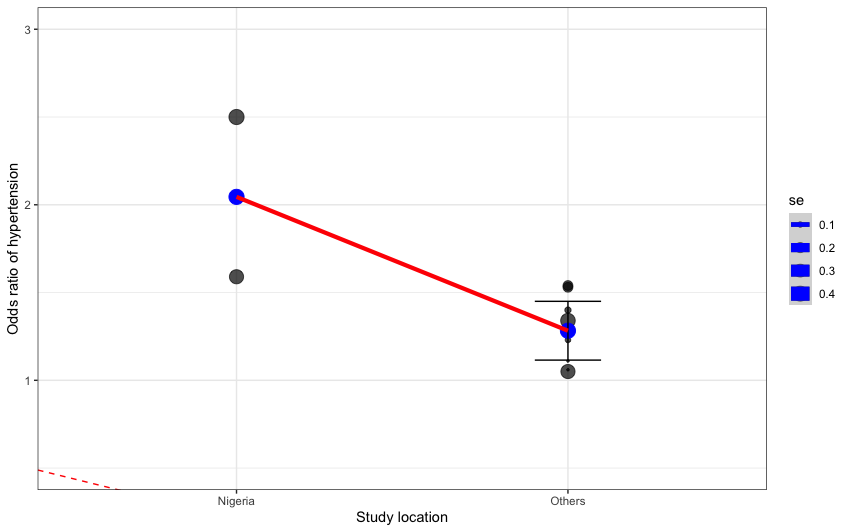

**(c)**

**Figure S6**. Bubble plot of 10 studies that reported on the association between dietary salt consumption and hypertension. The bubble plot illustrates the relationship of moderating effects of (A) mean age, (B) body mass index (BMI), and (C) study location on the association between dietary salt consumption and hypertension. The bubble plot in (A), (B), and (C) suggests that there is no significant moderating effect of mean age, BMI, and study location on the relationship between fruit consumption and hypertension.

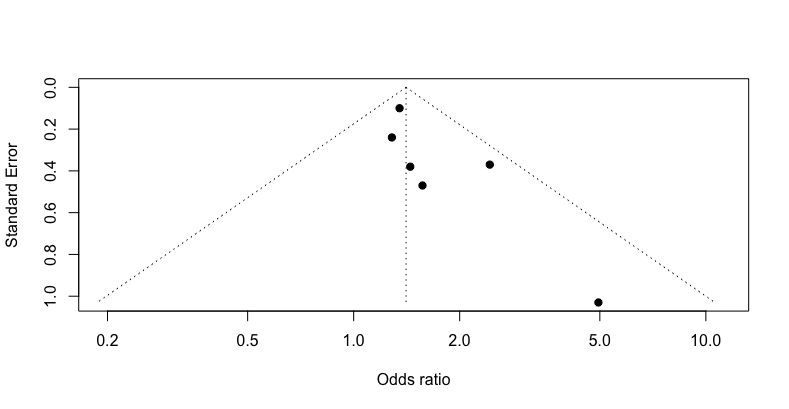

**Figure S7.** Funnel plots of 5 studies that reported on the association between junk food consumption and hypertension in West Africa with an odds ratio (x-axis) vs standard error (y-axis). No publication bias was demonstrated by the funnel plot, Egger’s regression test (p= 0.26), and rank correlation test (p= 0.06) in the meta-analysis.

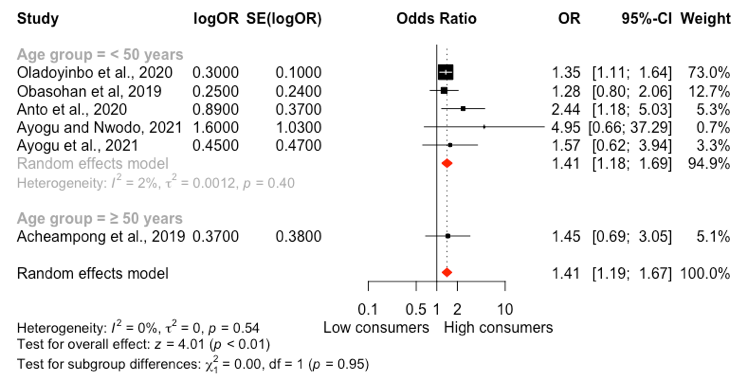

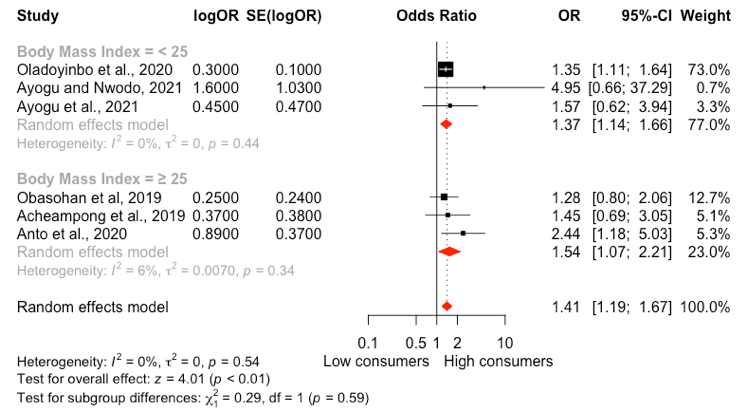

(A) **(B)**

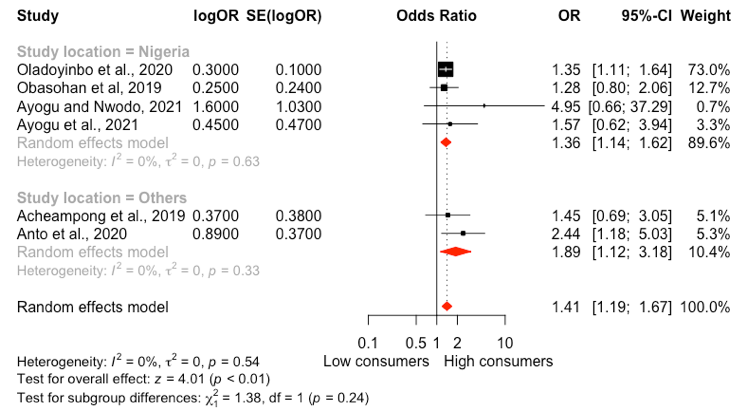

**(C)**

**Figure S8.** Forest plot of Subgroup analysis of the moderating effect of (A) mean age, (B) BMI, and study location (C) on the association between junk food consumption and hypertension in West Africa. No significant moderating effect was observed for mean age, BMI, and study location in (A), (B), and (C) on the effect size, respectively. CI: Confidence interval, logOR: Treatment effect, OR: Odds ratio, SE (logOR): Standard error.

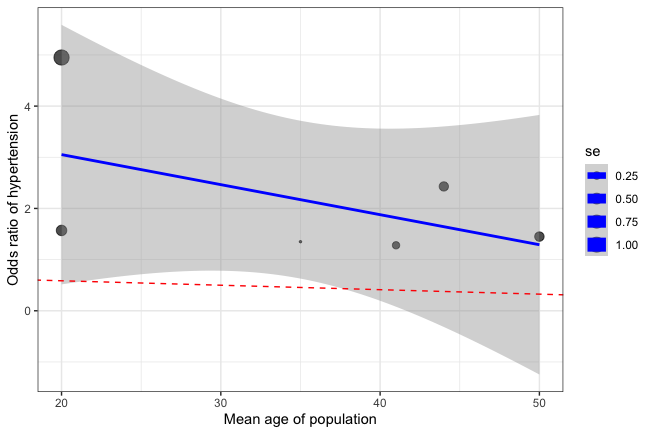

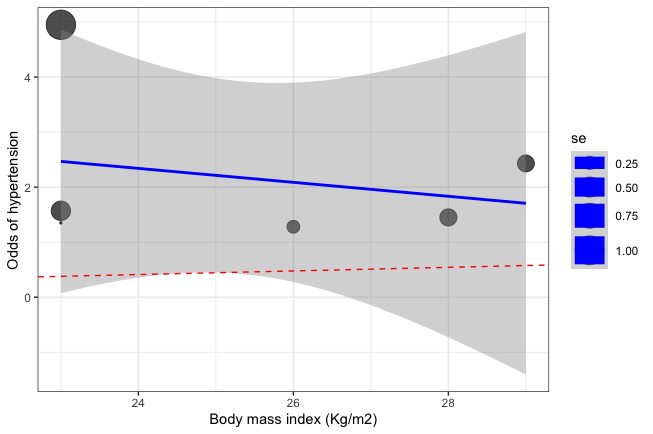

(A) **(B)**

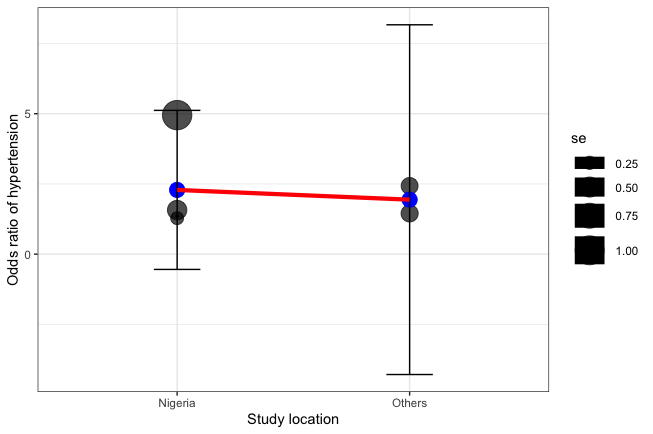

**(C)**

**Figure S9**. Bubble plot 6 studies that reported on the association between junk food consumption and hypertension. The bubble plot illustrates the relationship of moderating effects of (A) mean age, (B) body mass index (BMI), and (C) study location on the association between junk food consumption and hypertension. The bubble plot in (A), (B), and (C) suggests that there is no significant moderating effect of mean age, BMI, and study location on the relationship between fruit consumption and hypertension.

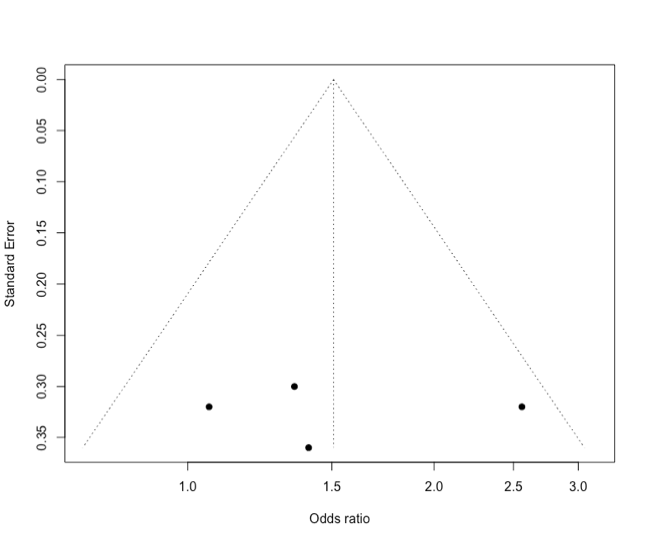

**Figure S10.** Funnel plots of 4 studies that reported on the association between red meat consumption and hypertension in West Africa with an odds ratio (x-axis) vs standard error (y-axis). No publication bias was indicated by the funnel plot, Egger’s regression test (p= 1.00), and rank correlation test (p= 0.72) in the meta-analysis.

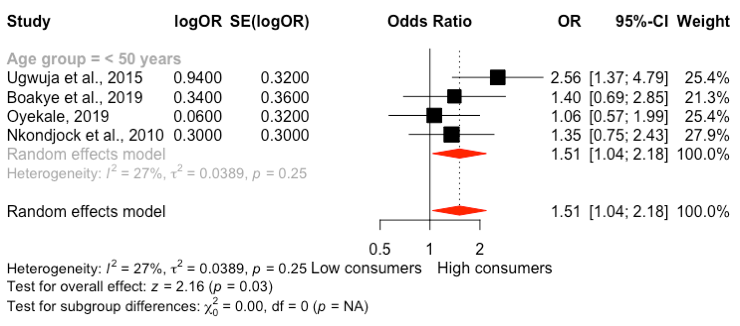

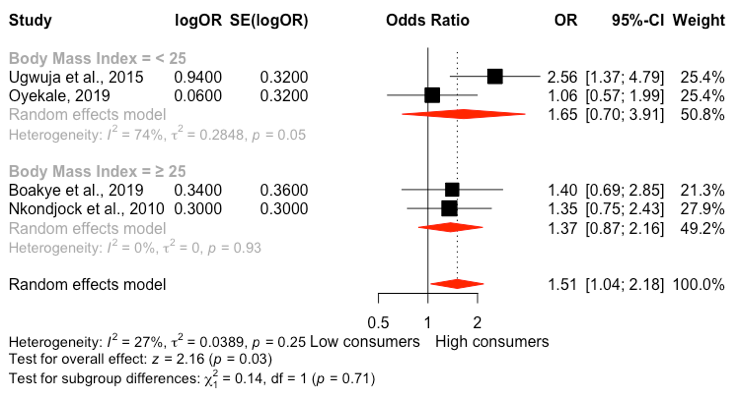

(A) **(B)**

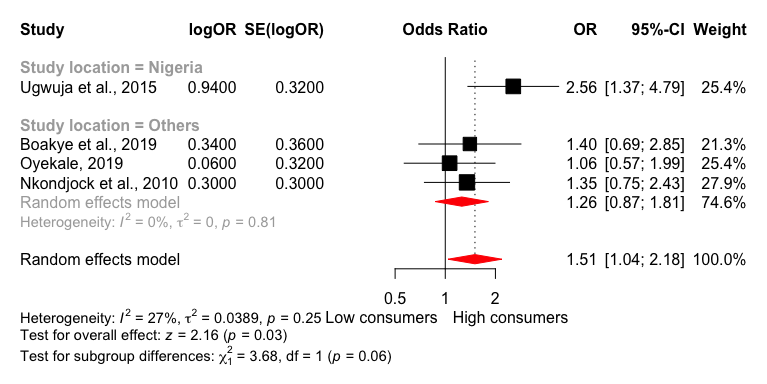

**(C)**

**Figure S11.** Forest plot of Subgroup analysis of the moderating effect of (A) mean age, (B) BMI, and study location (C) on the association between red meat consumption and hypertension in West Africa. No significant moderating effect was observed for mean age, BMI, and study location in figures (A), (B), and (C), respectively. CI: Confidence interval, logOR: Treatment effect, OR: Odds ratio, SE (logOR): Standard error.

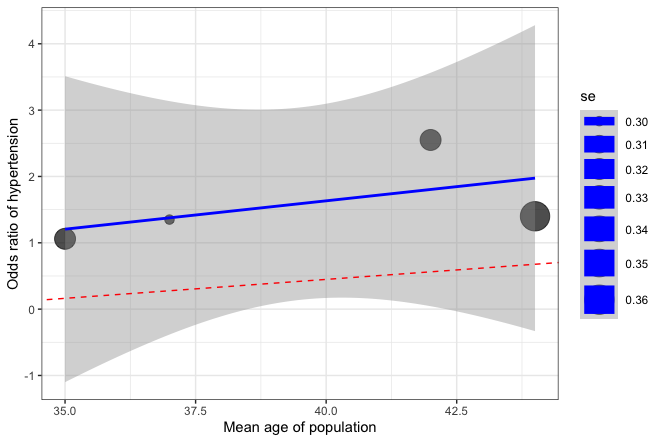

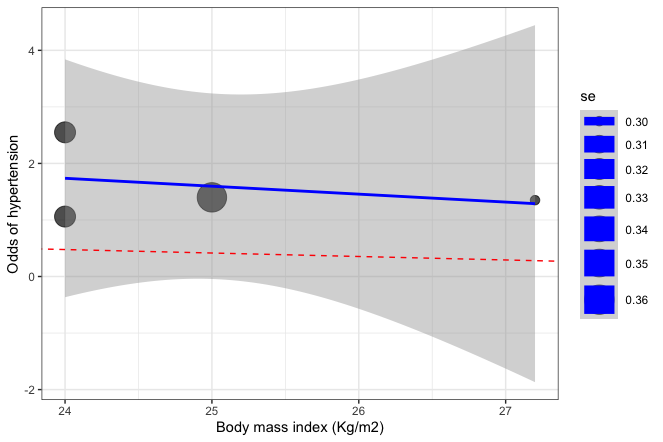

(A) **(B)**

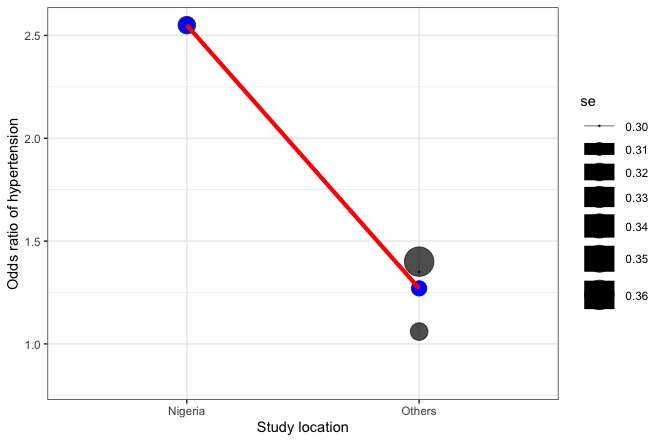

**(C)**

**Figure S12**. Bubble plot 4 studies that reported on the association between red meat consumption and hypertension. The bubble plot illustrates the relationship of moderating effects of (A) mean age, (B) body mass index (BMI), and (C) study location on the association between red meat consumption and hypertension. No moderating effect of the mean age, BMI, and study location on the relationship between red meat consumption and hypertension was observed.

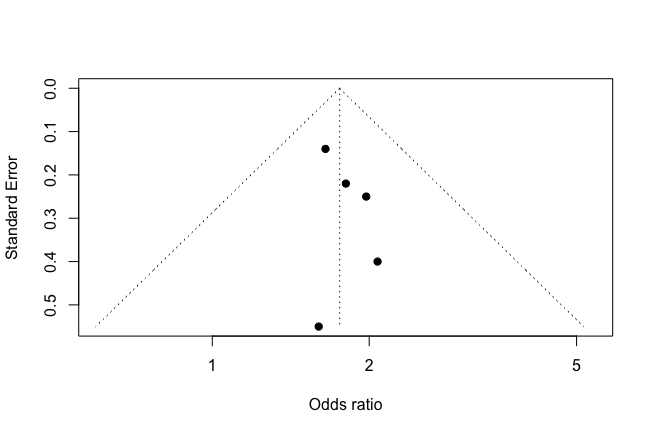

**Figure S13.** Funnel plots of 5 studies that reported on the association between dietary fat consumption and hypertension in West Africa with an odds ratio (x-axis) vs standard error (y-axis). No publication bias was demonstrated by the funnel plot, Egger’s regression test (p= 0.89), and Begg’s test (p= 0.48) in the meta-analysis.

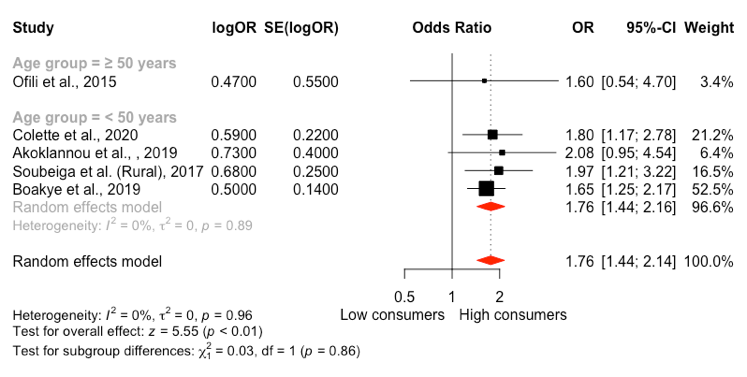

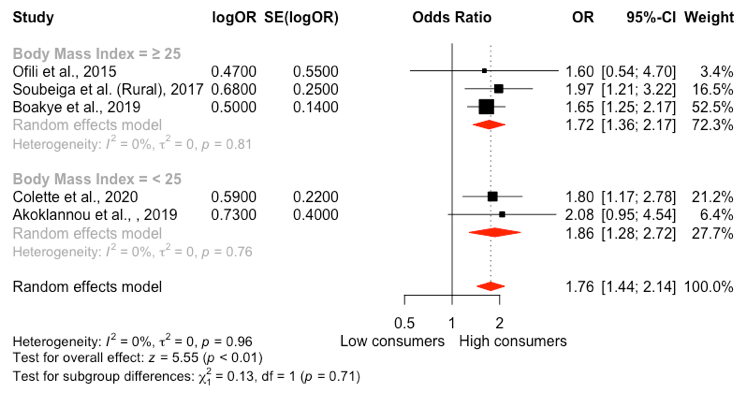

(A) **(B)**

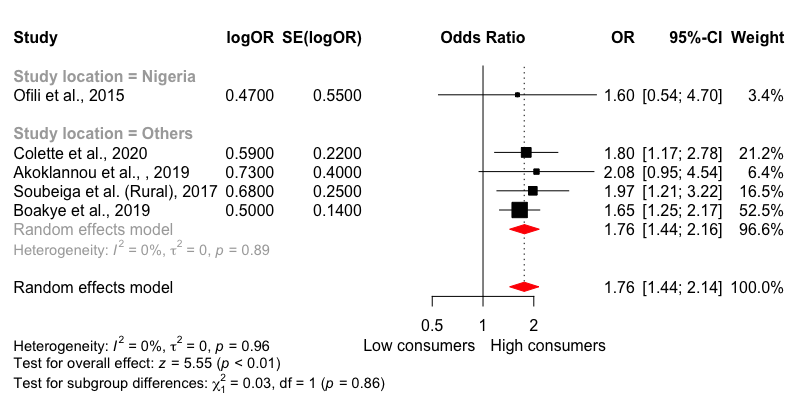

**(C)**

**Figure S14.** Forest plot of Subgroup analysis of the moderating effect of (A) mean age, (B) BMI, and study location (C) on the association between dietary fat consumption and hypertension in West Africa. No significant moderating effect was observed for mean age, BMI, and study location in figures (A), (B), and (C) on the effect size, respectively. CI: Confidence interval, logOR: Treatment effect, OR: Odds ratio, SE (logOR): Standard error.

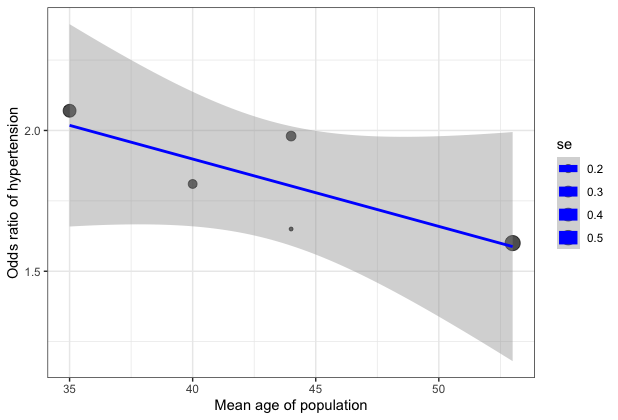

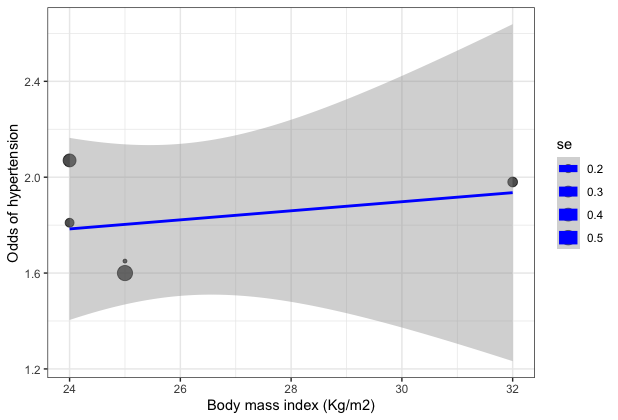

(A) **(B)**

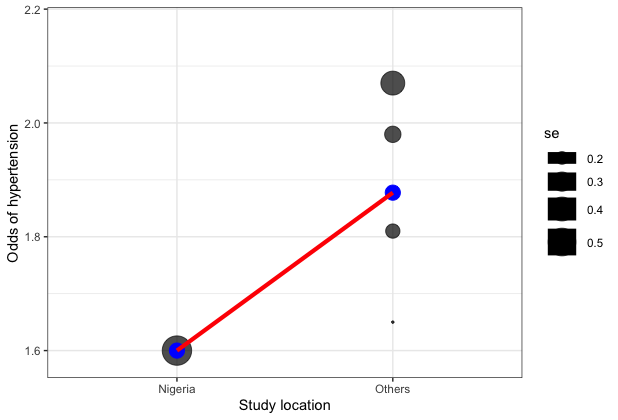

**(C)**

**Figure S15**. Bubble plot 5 studies that reported on the association between dietary fat consumption and hypertension. The bubble plot illustrates the relationship of moderating effects of (A) mean age, (B) body mass index (BMI), and (C) study location on the association between dietary fat consumption and hypertension. The bubble plot in (A), (B), and (C) suggests that there is no significant moderating effect of the mean age, BMI, and study location on the relationship between dietary fat consumption and hypertension.

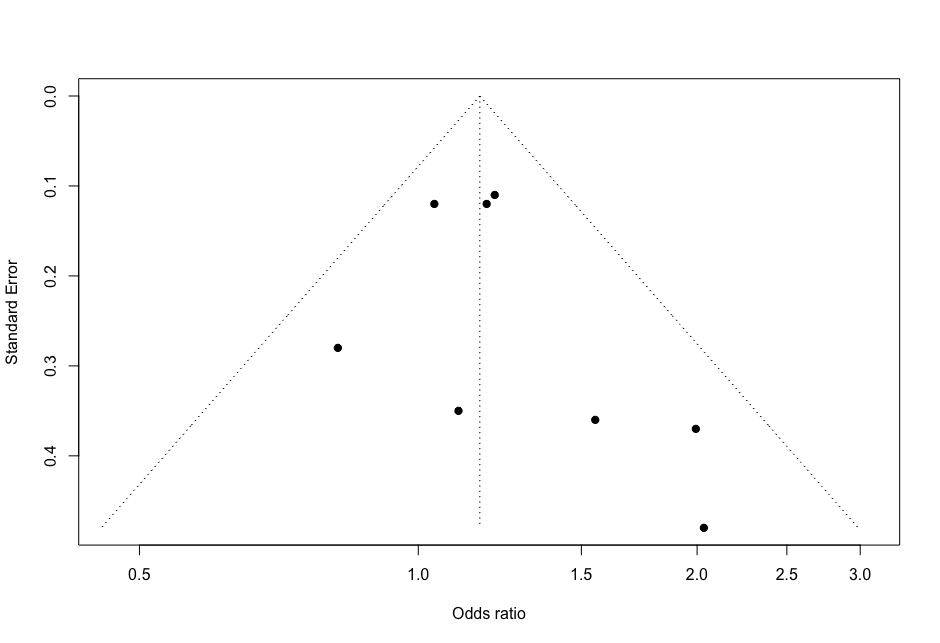

**Figure S16.** Funnel plots of 8 studies that reported on the association between alcohol consumption and hypertension in West Africa with an odds ratio (x-axis) vs standard error (y-axis). No publication bias was demonstrated by the funnel plot, Egger’s regression test (p= 0.46), and Begg’s test (p= 0.17) in the meta-analysis.

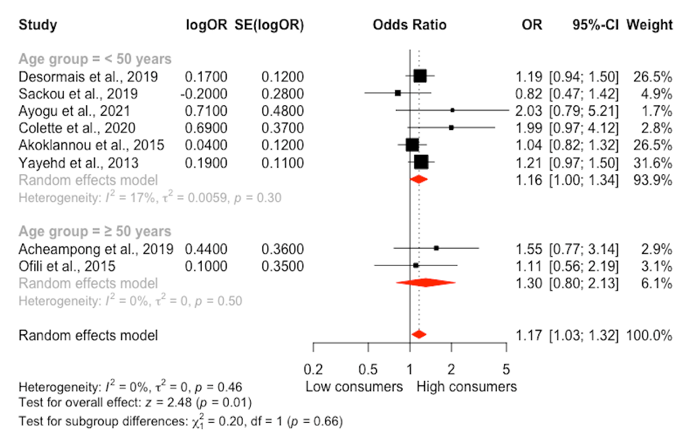

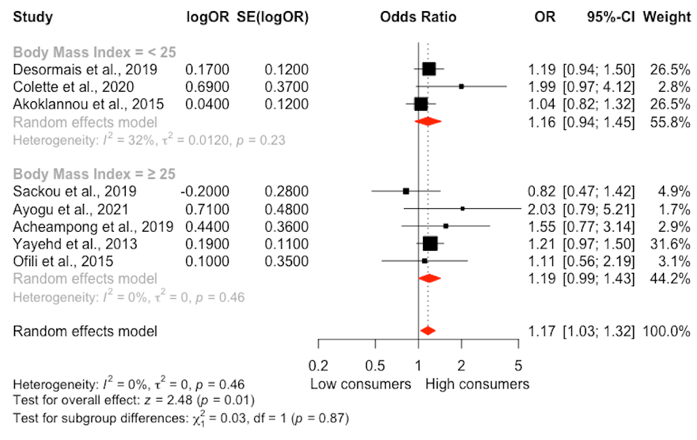

(A) (B)

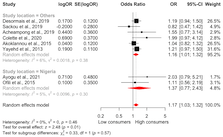

(C)

**Figure S17.** Forest plot of Subgroup analyses of the moderating effect of (A) mean age, (B) BMI, and study location (C) on the association between alcohol consumption and hypertension in West Africa. No significant moderating effect was observed for mean age, BMI, and study location in (A), (B), and (C) on the effect size, respectively. CI: Confidence interval, logOR: Treatment effect, OR: Odds ratio, SE (logOR): Standard error.

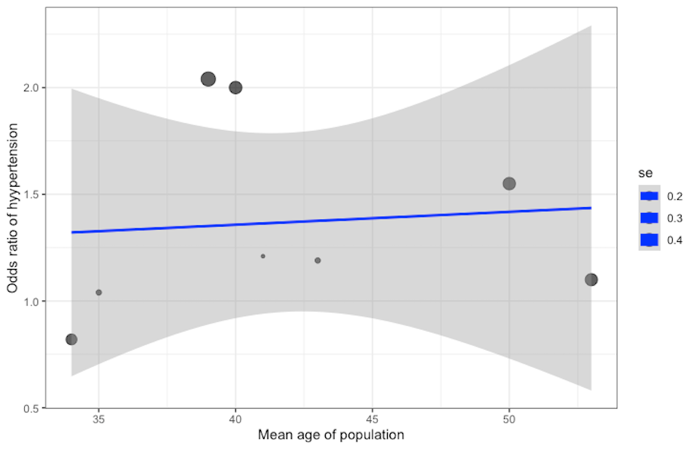

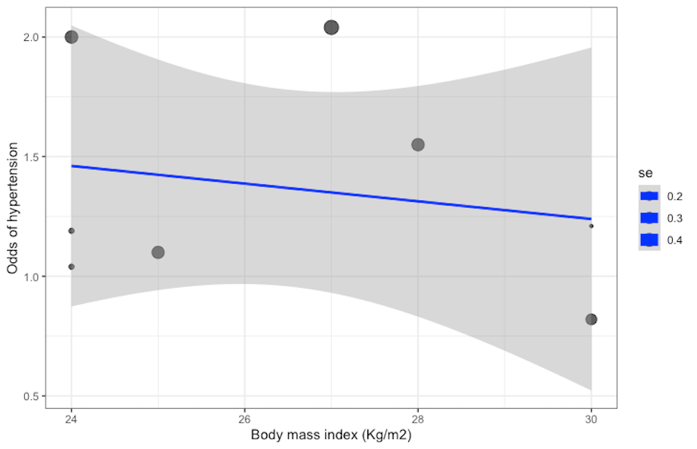

(A) **(B)**

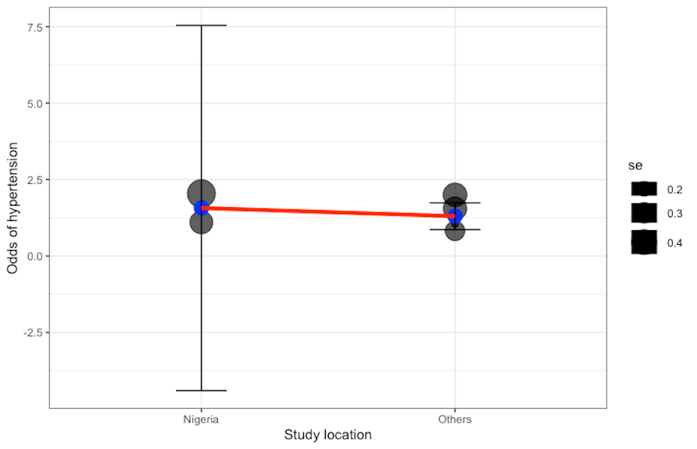

**(C)**

**Figure S18**. Bubble plot 5 studies that reported on the association between alcohol consumption and hypertension. The bubble plot illustrates the relationship of moderating effects of (A) mean age, (B) body mass index (BMI), and (C) study location on the association between alcohol consumption and hypertension. The bubble plot in (A), (B), and (C) suggests that there is no significant moderating effect of the mean age, BMI, and study location on the relationship between alcohol consumption and hypertension.

**Table S4:** PRISMA-P (Preferred Reporting Items for Systematic review and Meta-Analysis Protocols) checklist: for dietary factors and hypertension in West Africa.

| Section and topic | Item No | Checklist item | Reported on page # |
| --- | --- | --- | --- |
| **TITLE** |  |  |  |
| Identification | 1 | Identify the report as a protocol of a systematic review and meta-analysis | **1** |
| **AUTHORS** |  |  |  |
| Contact | 2 | Provide name, institutional affiliation, e-mail address of all protocol authors; provide physical mailing address of corresponding author | **1** |
| **ABSTRACT** |  |  |  |
| Structured summary | 3 | Provide a structured summary including, as applicable: background; objectives; data sources; study eligibility criteria, participants, and interventions; study appraisal and synthesis methods; results; limitations; conclusions and implications of key findings; systematic review registration number | **2** |
| INTRODUCTION | | |  |
| Rationale | 5 | Describe the rationale for the review in the context of what is already known | **3** |
| Objectives | 6 | Provide an explicit statement of the question(s) the review will address with reference to participants, interventions, comparators, and outcomes (PICO) | **3** |
| METHODS | | |  |
| Protocol and registration | 7 | Indicate if a review protocol exists, if and where it can be accessed (e.g., Web address), and, if available, provide registration information including registration number. | **5** |
| Eligibility criteria | 8 | Specify the study characteristics (such as PICO, study design, setting, time frame) and report characteristics (such as years considered, language, publication status) to be used as criteria for eligibility for the review | **6** |
| Information sources | 9 | Describe all intended information sources (such as electronic databases, contact with study authors, trial registers or other grey literature sources) with planned dates of coverage | **5** |
| Search strategy | 10 | Present draft of search strategy to be used for at least one electronic database, including planned limits, such that it could be repeated | **5, Table S1** |
| Data management | 11 | Describe the mechanism(s) that will be used to manage records and data throughout the review | **6** |
| Study selection process | 12 | State the process that will be used for selecting studies (such as two independent reviewers) through each phase of the review (that is, screening, eligibility and inclusion in meta-analysis) | **6** |
| Data collection process | 13 | Describe planned method of extracting data from reports (such as piloting forms, done independently, in duplicate), any processes for obtaining and confirming data from investigators | **6** |
| Data items | 14 | List and define all variables for which data will be sought (such as PICO items, funding sources), any pre-planned data assumptions and simplifications | **5** |
| Risk of bias in individual studies | 15 | Describe anticipated methods for assessing risk of bias of individual studies, including whether this will be done at the outcome or study level, or both; state how this information will be used in data synthesis | **7** |
| Summary measures | 16 | State the principal summary measures (e.g., risk ratio, difference in means). | **6** |
| Synthesis of results | 17 | Describe the methods of handling data and combining results of studies, if done, including measures of consistency (e.g., I2) for each meta-analysis. | **7** |
| Risk of bias across studies | 18 | Specify any assessment of risk of bias that may affect the cumulative evidence (e.g., publication bias, selective reporting within studies). | **7** |
| Additional analyses | 19 | Describe methods of additional analyses (e.g., sensitivity or subgroup analyses, meta-regression), if done, indicating which were pre-specified. | **8** |
| Data synthesis | 20 | Describe criteria under which study data will be quantitatively synthesised.  If data are appropriate for quantitative synthesis, describe planned summary measures, methods of handling data and methods of combining data from studies, including any planned exploration of consistency (such as I^2^, Kendall’s τ)  Describe any proposed additional analyses (such as sensitivity or subgroup analyses, meta-regression)  If quantitative synthesis is not appropriate, describe the type of summary planned | **8** |
| **RESULTS** | | |  |
| Literature search/study selection | 21 | Give numbers of studies screened, assessed for eligibility, and included in the review, with reasons for exclusions at each stage, ideally with a flow diagram. | **9, Figure 1** |
| Study characteristics | 22 | For each study, present characteristics for which data were extracted (e.g., study size, PICOS, follow-up period) and provide the citations. | **9** |
| Risk of bias within studies | 23 | Present data on risk of bias of each study and, if available, any outcome level assessment (see item 12). | **9** |
| Results of individual studies | 24 | For all outcomes considered (benefits or harms), present, for each study: (a) simple summary data for each intervention group (b) effect estimates and confidence intervals, ideally with a forest plot. | **Table S2** |
| Synthesis of results | 25 | Present results of each meta-analysis done, including confidence intervals and measures of consistency. | **12-30** |
| Studies quality | 26 | Present results of any assessment of risk of bias across studies (see Item 15). | **Table S3** |
| Additional analysis | 27 | Give results of additional analyses, if done (e.g., sensitivity or subgroup analyses, meta-regression [see Item 16]). | **Figure S3- 18** |
| **DISCUSSION** | | |  |
| Summary of evidence | 28 | Summarize the main findings including the strength of evidence for each main outcome; consider their relevance to key groups (e.g., healthcare providers, users, and policy makers). | **31-38** |
| Limitations | 29 | Discuss limitations at study and outcome level (e.g., risk of bias), and at review-level (e.g., incomplete retrieval of identified research, reporting bias). | **38** |
| Conclusions | 30 | Provide a general interpretation of the results in the context of other evidence, and implications for future research. | **38** |
| **SUPPORT** | | | |
| Sponsor | 4 | Provide name for the review funder and/or sponsor | **39** |
